# A cognitive behavioral therapy program designed for low-literacy end-users with perinatal depression in Sierra Leone: an investigator- and outcomes assessor-blinded, controlled randomized trial

**DOI:** 10.1101/2025.09.29.25336811

**Authors:** Eliza Kleban, Ashleen Lee, Aminata Koroma, D. Taylor Hendrixson, Joshua Duncan, Kevin B Stephenson, Mark J Manary

**Affiliations:** School of Medicine, Washington University, St. Louis, MO, USA; Project Peanut Butter, Freetown, Sierra Leone; Ministry of Health, Government of Sierra Leone, Freetown, Sierra Leone; Department of Pediatrics, University of Washington, Seattle, WA USA; Mental Health Coalition-Sierra Leone, Freetown, Sierra Leone; Children’s Nutrition Research Center, Baylor College of Medicine, Houston, TX, USA

## Abstract

**Background:** Perinatal depression is common in Sub-Saharan Africa, where mental health services are limited and literacy levels low. Cognitive behavioral therapy (CBT) is effective in high-income settings, but uses written materials. We evaluated the effectiveness of a culturally adapted CBT intervention designed for non-literate women with perinatal depression, delivered by trained lay counselors in rural Sierra Leone.

**Methods:** This study was an outcomes assessor- and investigator-blinded, individually randomized controlled trial of an adapted CBT program versus standard care (no mental health intervention) among undernourished pregnant and postpartum women with depression in Pujehun, Sierra Leone. Participants were drawn from the COGENT cohort, a randomized, controlled trial of antenatal nutritional supplementation. Depression was identified using the adapted Patient Health Questionnaire-9 (aPHQ-9). Eligible women scoring ≥9 on the aPHQ-9 were randomized 1:1 to receive six weekly CBT sessions or standard care. The primary outcome was aPHQ-9 score at 8 weeks post-randomization. Secondary outcomes included clinically meaningful symptom reduction and depression remission (aPHQ-9 <5). Outcomes were analyzed using a modified intention-to-treat analyses.

**Findings:** Among the 881 women screened, 155 were randomized to CBT i (n=80) or control (n=75) groups. Endline data were available for 140/ 153 participants (92%). Median aPHQ-9 scores at 8 weeks were lower in the CBT group than the control group (median difference −4, 95% CI −5 to −3; p<0.001). Clinically symptom reductions (>3-point drop) were more common in the CBT group (96% vs 55%; OR 19, 95% CI 6 to 86; p<0.001), and 79% of CBT participants achieved remission compared to 34% of controls (OR 7, 95% CI 4 to 16; p<0.001). Effects were sustained through 9 months postpartum. No significant effect heterogeneity was found across subgroups. Inter-assessor reliability of aPHQ-9 scoring was high (ICC >0.90).

**Interpretation:** A culturally adapted CBT program reduced perinatal depression symptoms in malnourished antenatal and postpartum women in rural Sierra Leone, with sustained effects through nine months postpartum. This study demonstrates the feasibility and effectiveness of delivering CBT via trained lay counselors. Funding provided by Open Philanthropy.

## INTRODUCTION

Perinatal mood disorders encompass a spectrum of mental health conditions occurring during pregnancy and up to one year postpartum, including perinatal depression, anxiety disorders, and in severe cases, perinatal psychosis. Perinatal depression in particular represents a major global health challenge, with prevalence rates of 10-20% in high-income countries and substantially higher rates reported in low- and middle-income countries.^1^ In sub-Saharan Africa, the burden is particularly pronounced, with studies reporting perinatal depression rates ranging from 15-40%, influenced by factors such as poverty, social isolation, intimate partner violence, HIV infection, and limited social support systems.^2–4^ Perinatal depression is associated with poor self-care, impaired mother-infant bonding, preterm birth, low birth weight, and impaired offspring physical and neurodevelopment. Despite the availability of effective treatments, access to mental health care remains severely limited in resource-constrained settings, with treatment gaps exceeding 90% in many sub-Saharan African countries, where cultural stigma around mental health, shortage of trained mental health professionals, and competing healthcare priorities further compound the challenge.

Cognitive behavioral therapy (CBT) has demonstrated efficacy for treating perinatal depression in multiple randomized, controlled trials. However, most evidence comes from high-income settings with literate populations. Standard CBT protocols rely heavily on written materials, homework assignments, and literacy-dependent exercises that may not be feasible or culturally appropriate in low-resource, low-literacy contexts.

Community based delivery approaches using trained non-specialist counselors have shown promise for delivering mental health interventions in resource-limited settings.^5–7^ However, few rigorously evaluated programs exist specifically for perinatal depression in rural African settings, and none have been tested among populations with very low literacy rates. Adaptation requires not only linguistic translation but fundamental redesign of therapeutic approaches to function without written materials while maintaining core CBT principles.

We previously developed a CBT intervention adapted for pregnant and postpartum women with depression and low literacy in Pujehun District, Sierra Leone, a setting with no pre-existing mental health services and a female literacy rate below 15%. We aimed to evaluate the effectiveness of this adapted CBT program on improving perinatal depression compared to standard care among undernourished pregnant and postpartum women with depression.

## METHODS

### Study Design and Setting

We conducted an individually randomized, controlled, outcomes assessor- and investigator-blinded superiority trial (hereafter referred to as CBT Trial) of a CBT program developed for low-literacy pregnant and postpartum women with depression versus no treatment as part of a 2×2 factorial trial in Pujehun District, Sierra Leone, between 2023 and 2025. In this setting, “no treatment” reflects the standard of care, which does not currently include formal mental health services. The parent trial, “Improving COgnition and GEstational duration with targeted NuTrition” (COGENT), simultaneously tested antenatal supplementation with a ready-to-use supplementary food with or without added omega-3 fatty acids and choline. COGENT and CBT Trial randomizations were independent. All CBT Trial participants were randomized within COGENT, but only a subset of COGENT participants were enrolled in the CBT Trial.

The CBT trial was conducted at six government-run antenatal clinics selected from 12 COGENT sites based on larger catchment populations in Pujehun District, Sierra Leone. Pujehun is a rural district in southeastern Sierra Leone with no electrical grid, high rates of undernutrition in pregnancy, low literacy rates, and no existing mental health interventions for perinatal depression at the time of study initiation.

The study protocol was approved by the Sierra Leone Ethics and Scientific Review Committee, the Pharmacy Board of Sierra Leone, and the Washington University Human Research Protection Office. The trial is registered at ClinicalTrials.gov (NCT05949190). The full protocol is available in the supplementary appendix (Appendix 1). The trial was conducted in compliance with the International Council for Harmonization guidelines for good clinical practice, including local monitoring visits and central data monitoring.

### Participants

CBT Trial participants were pregnant and postpartum women enrolled in COGENT who developed depression during COGENT study participation. All COGENT participants underwent depression screening using an adapted Patient Health Questionnaire-9 (aPHQ-9) at enrollment, monthly during pregnancy, and at 1.5, 3, 6, and 9 months postpartum. Women at the 6 selected clinics scoring ≥ 9 on the aPHQ-9 at any screening point were eligible for the CBT trial. Participants were also encouraged to present any time they felt concerned about their mental or physical health.

COGENT eligibility criteria were: pregnant, aged ≥13 years, undernourished as defined by mid-upper arm circumference ≤23.0 cm and/or BMI <18.5, willingness to comply with study procedures, and planning to reside in the study area. Exclusion criteria included participation in other supplementary feeding programs, known allergies to study foods or medications, and conditions requiring immediate hospitalization. There were no additional exclusion criteria for the CBT Trial. For participants under 18 years who were unmarried, both participant assent and parent/guardian consent were required. Participants provided written informed consent or thumbprint consent with an impartial witness present for those unable to read.

### Randomization and masking

Participants were individually randomized 1:1 to CBT intervention or control using an opaque-envelope manual randomization system. A research staff member not otherwise involved in the trial prepared blocks of 24 small opaque envelopes, each containing one of six symbols, with four of each symbol per block. Three symbols were assigned to each arm. Each block of 24 was placed in a larger, opaque envelope. Participants blindly selected a smaller envelope to determine allocation.

Given the nature of the intervention, participants and treating counselors could not be blinded. Outcome assessors and investigators, including the trial statistician, were blinded to allocation. At each clinic, the counselor who conducted screening, enrollment, and randomization delivered CBT if allocated, while a different counselor was assigned to conduct outcome assessments.

### Study Procedures

The CBT program was developed in 2022-2023 in collaboration with the Mental Health Coalition-Sierra Leone (MHC-SL). Counselors were hired based on female sex, Pujehun residency, fluency in the local dialect, Mende, prior experience with perinatal depression, and observed ease establishing rapport with clients. CBT was then adapted for use by clients without literacy. Thought records and written homework were replaced with oral processing, mental rehearsal and role-play, and drawing. Pictorial aids were used to complement symptom assessment. MHC-SL trained counselors on identification of depression and implementation of this adapted CBT program, with monthly direct oversight visits to assess quality and fidelity. Program development is described in detail elsewhere.^8^

The CBT intervention consisted of six weekly 45–60-minute individual sessions. Sessions followed a structured protocol: (1) problem identification and psychoeducation; (2-3) solution generation and selection using SMART (Specific, Measurable, Attainable, Realistic, Timely) goals; (4-5) evaluation of attempted solutions and skill refinement; (6) integration and relapse prevention. Between sessions, participants received tailored homework assignments practiced with counselors to assure comprehension. Sessions occurred primarily at clinics, with occasional home visits when transportation was not feasible. Full details of the intervention have been previously published.^8^

Control group participants received no specific mental health intervention as is current standard of care in this setting but continued to receive all standard care available through the COGENT study, including routine antenatal and postnatal care, and antenatal nutritional supplementation.

Safety oversight was provided by MHC-SL experts, who were available for immediate referral for aPHQ-9 scores ≥ 20 or reported suicidality, and the COGENT study’s Local monitor and Independent Medical Reviewer, in accordance with the Pharmacy Board of Sierra Leone’s guidelines.

### Outcomes

The primary outcome was aPHQ-9 score at 8 weeks post-randomization. The PHQ-9 was selected for adaptation based on its widespread use and validation in sub-Saharan Africa, including Ethiopia, Kenya, and Nigeria.^9–11^ More recently, aPHQ-9 was validated in Sierra Leone, demonstrating a sensitivity of 73.8%, specificity of 76.2%, and an internal consistency reliability of 0.81.^12^ The aPHQ-9 differs from the PHQ-9 in 4 ways: (1) translation to the local language, Mende, (2) addition of visual aids to help illustrate symptom frequency, (3) question 5 reference to overeating was removed, as all participants were undernourished, and (4) question 7 “reading newspaper or watching television” was replaced with “doing household chores”.^8^ These changes were made in consultation with MHC-SL to improve cultural relevance. Translations were iteratively improved with field testing prior to implementation in the CBT Trial.^8^ Like the PHQ-9, the aPHQ-9 consists of nine items scored 0 (“not at all”) to 3 (“nearly every day”), for a total score of 0-27. Among participants who missed their 8-week outcome assessment visit, aPHQ-9 scores collected up to 16 weeks post-randomization were included.

Secondary outcomes were change in aPHQ-9 between baseline and endline, reduction in aPHQ-9 between baseline and endline of ≤ 3, >3, >5, or >9 points, reduction between baseline and endline of >50%, endline aPHQ-9 <5, as well as aPHQ-9 scores measured after endline assessment as part of routine data collection in COGENT. Inter-assessor reliability of aPHQ-9 assessment was evaluated by having two different counselors assess aPHQ-9 scores on a single participant one hour apart. All 4 counselors underwent inter-assessor reliability testing during two separate weeks of the trial.

### Sample Size & Statistical analysis

The study planned to enroll 75 women per arm for a total of 150. Anticipating 10% loss-to-follow-up, this sample size provided 80% power and a two-sided α=0.05 to detect a 2-point aPHQ-9 difference, corresponding to a Cohen’s d=0.5 at SD=4. This SD was anticipated based on pilot data from Pujehun District.^8^ A 2-point difference is in the range of those detected in prior trials of CBT for peripartum depression (citations).

Data were double entered from paper forms into password-protected Microsoft Access databases and verified for accuracy.

Analyses were conducted in two populations: a modified intention-to-treat population which included all participants correctly enrolled and with primary outcome endline aPHQ-9 data, and a modified intention-to-treat population that included all participants correctly enrolled. Participants who were subsequently found not to have been pregnant at the time of randomization were excluded. Participants were analyzed according to their allocated group irrespective of treatment received or adherence to it. For the primary outcome, the Wilcoxon rank-sum test was used to compare aPHQ-9 scores. The Hodges-Lehmann estimator was used to compute the median of differences between groups.^13^ A 95% CI for the median of differences was estimated using the normal approximation method with continuity correction, to account for frequent ties expected in aPHQ-9 scores. For binary outcomes, ORs and 95% CIs were estimated using logistic regression.

In two post-hoc sensitivity analyses, participants without endline aPHQ-9 data had values imputed. In one scenario, assuming poor results among those with missing data, endline aPHQ-9 scores were imputed by adding 2 points to baseline scores. In an alternative, better-case scenario, 9 points were subtracted from baseline scores to impute endline scores. We conducted best- and worst-case imputation scenarios using fixed offsets to baseline scores, as justified in prior methodological work.^14,15^

Pre-specified secondary analyses adjusted for age, education level, and ante- vs. postnatal status were done for continuous outcomes using ordinal logistic regression and for binary outcomes using logistic regression. The proportional odds assumption was deemed met after plotting empirical logits against cut-point values for each treatment group and identifying parallelism (Harrell). Using ordinal logistic regression, we assessed for heterogeneity of effect in endline aPHQ-9 scores by plotting ORs and 95% CIs by subgroup as well as estimating a p value for the interaction term between study group and each subgroup. In all cases, OR>1 indicates greater odds for a lower aPHQ-9 score. Pre-specified subgroups were maternal age, education level, randomized COGENT nutrition supplement group, and baseline aPHQ-9; post-hoc subgroups were first pregnancy vs. other, father in home, and gestational age at CBT enrollment among participants enrolled antenatally. Inter-assessor reliability of aPHQ-9 assessment was analyzed using a two-way random-effects model and reported as ICC (95% CI). All statistical analyses were performed using R version 4.5.0.

## RESULTS

Between November 8, 2023 and April 28, 2025, 881 women were screened for eligibility and 155 participants were randomly assigned to the CBT group (n = 80) or control group (n = 75) (Figure 1).

**Figure 1.**
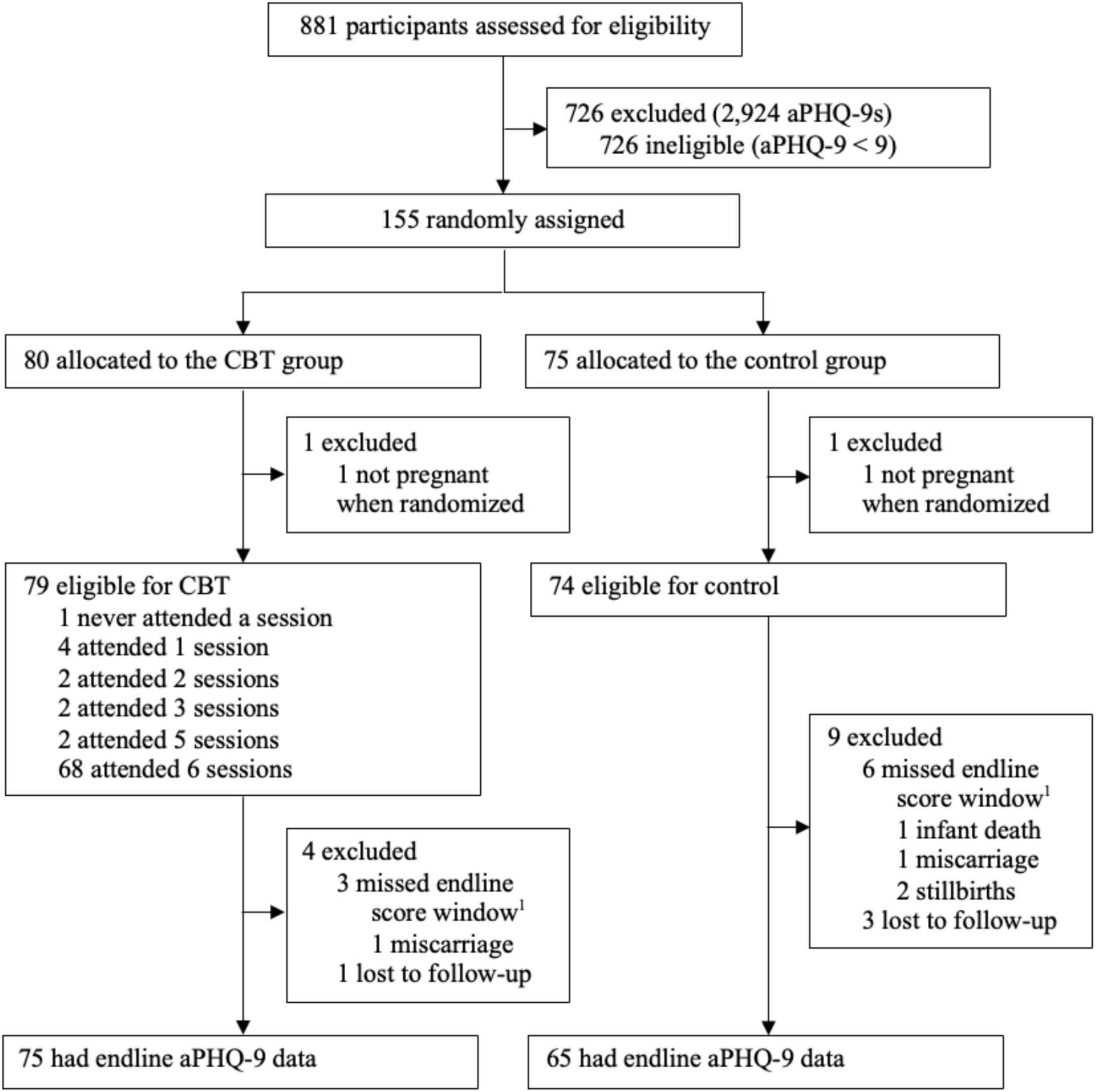
Trial profile. CBT=cognitive behavioral therapy. mITT=modified intention-to-treat. * When available, reasons provided for missing endline assessment window are indicated.

One participant in each group was then found not to have been pregnant at the time of randomization, and both participants were excluded. Baseline characteristics of participants were similar between groups (Table 1). Among all participants, the median age was 19 years (IQR: 18, 22 years), and 134 (88%) out of 153 were enrolled during the antenatal period (Table 1). 35 (23%) of 153 participants reported no formal education, and 100 (65%) of 153 reported having completed secondary education. The reported rate of literacy was 37% (36/98). The median aPHQ-9 score at enrollment was 11 in both groups. Endline aPHQ-9 data were collected in 75 (94%) of 79 CBT participants and 65 (87%) of 74 control participants. With only 13 (8%) of 153 participants missing endline aPHQ-9 data, comparison of baseline characteristics among those with vs. without endline data was limited, revealing that control participants without primary outcome data were slightly older and had lower baseline aPHQ-9 scores, but were otherwise similar (Appendix 2). Among CBT participants, 68/ 79 (86%) attended all 6 counseling sessions.

**Table 1.** Selected baseline participant characteristics ^1^.

| Characteristic | CBT<br>(n = 79) | Control<br>(n = 74) |
| --- | --- | --- |
| Age, years, median (IQR)<sup>2</sup> | 19 (18, 22) | 19 (18, 23) |
| MUAC at enrollment, cm, median (IQR) | 22.5 (21.8, 23.0) | 22.4 (21.4, 22.9) |
| Weeks in supplementary feeding trial before enrollment in this trial, median (IQR)<sup>3</sup> | 4 (0, 10) | 4 (0, 10) |
| 0 | 37 (47) | 33 (45) |
| ≤ 4 | 14 (18) | 11 (14) |
| > 4 | 28 (35) | 30 (41) |
| Pregnancy status at enrollment |  |  |
| Antenatal | 70 (89) | 64 (86) |
| Gestational age at enrollment, weeks, median (IQR)<sup>4</sup> | 22.5 (18.9, 28.7) | 21.2 (17.9, 26.6) |
| Postnatal | 9 (11) | 10 (14) |
| Weeks since delivery at enrollment | 13.7 (12.5, 19.0) | 6.4 (6.0, 6.9) |
| Mother's education level |  |  |
| None | 14 (18) | 21 (28) |
| Primary | 7 (9) | 11 (15) |
| Secondary or greater | 58 (73) | 42 (57) |
| Currently in school<sup>5</sup> | 32 (41) | 16 (22) |
| History of miscarriage or stillbirth | 2 (3) | 7 (10) |
| Number of adults in household, median (IQR) | 4 (3, 5) | 4 (3, 5) |
| Two or more children live in household | 58 (73) | 58 (78) |
| Father lives in household | 54 (68) | 47 (64) |
| Household roof made of thatch | 10 (13) | 8 (11) |
| Pit latrine used for stool disposal | 67 (85) | 61 (82) |
| Household has electricity | 7 (9) | 11 (15) |
| Household Food Insecurity Access Scale score (past 4 weeks) |  |  |
| Food secure | 2 (2) | 8 (11) |
| Mildly food insecure | 0 (0) | 1 (1) |
| Moderately food insecure | 32 (41) | 21 (28) |
| Severely food insecure | 45 (57) | 44 (60) |
| aPHQ-9 score at time of enrollment, median (IQR) | 11 (10, 12) | 11 (9, 12) |
| Score of 9 | 11 (14) | 20 (28) |
| Score 10-12 | 52 (66) | 43 (59) |
| Score ≥ 13 | 16 (20) | 9 (13) |

<sup>1</sup> Values are n (%) unless otherwise indicated<sup>2</sup> n = 2 missing from CBT; n = 1 missing from control<sup>3</sup> CBT trial participants are a subset of a larger supplementary feeding study, details of which can be found in the Appendix.<sup>4</sup> n = 2 missing from control due to miscarriage prior to gestational age estimation being possible<sup>5</sup> n = 1 missing from control

In the primary mITT analysis population including only those with endline aPHQ-9 scores, aPHQ-9 scores were significantly lower in the CBT group compared to control group, with a median of differences of −4 (95% CI −5, −3; p<0.001) (Table 2, Figure 2). CBT participants achieved significantly higher rates of clinically meaningful symptom reduction across all predefined thresholds, with 96% achieving >3-point reduction compared to 55% of controls (OR 19, 95% CI 6, 86; p < 0.001). Additionally, 79% of CBT participants achieved remission (aPHQ-9 < 5) compared with 34% of control participants (OR 7, 95% CI 4, 16; p < 0.001) (Table 2). Among participants enrolled antenatally, aPHQ-9 scores were 2 points lower in the CBT group postpartum at 1.5 months (95% CI −3, 0; p = 0.007), 1 point lower at 3 months (95% CI −3, 0; p = 0.01), and 1 points lower at 9 months (95% CI −2, 0; p = 0.047), while scores were not different at 6 months postpartum.

**Figure 2.**
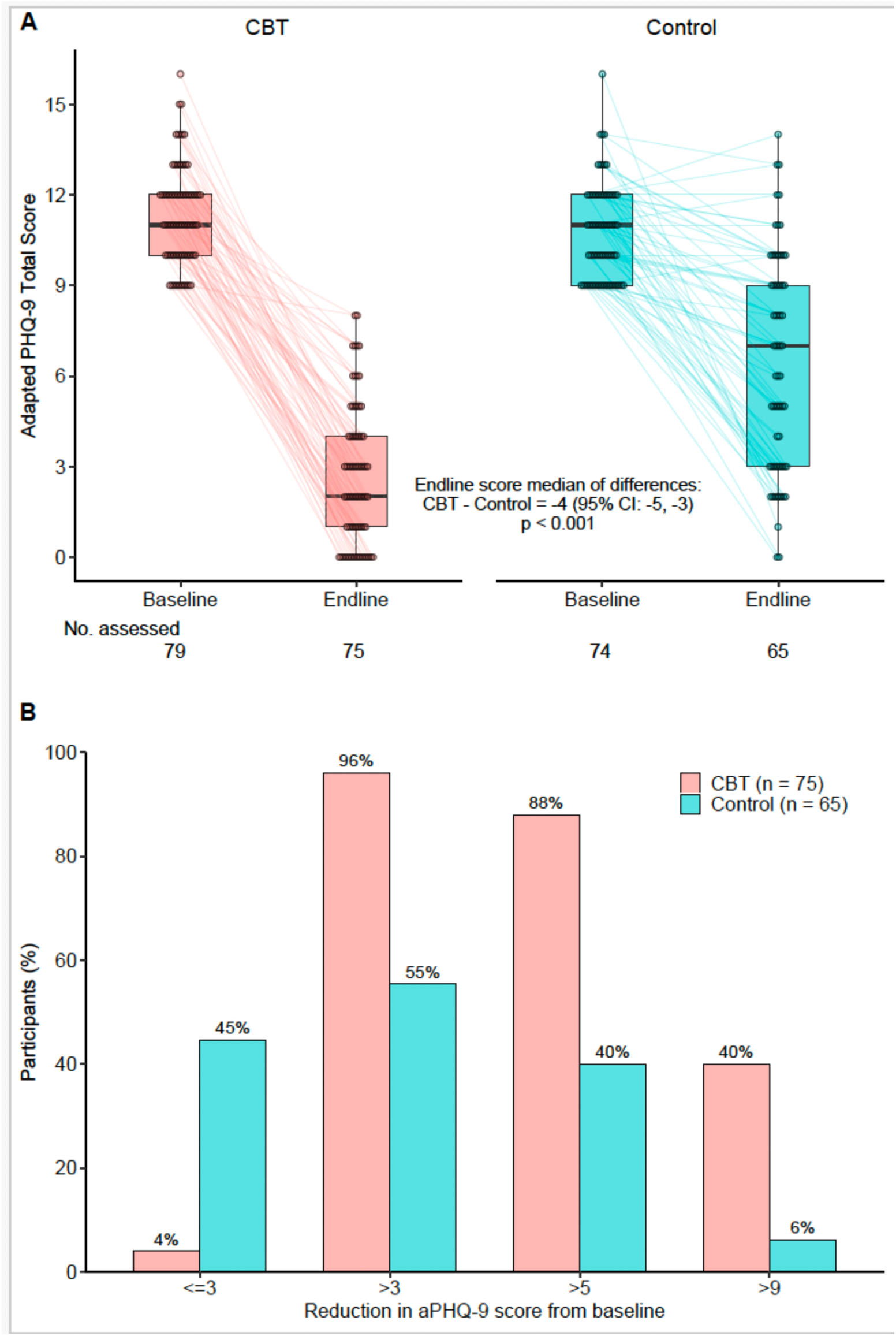
Effect of CBT program on adapted Patient Health Questionnaire-9 scores. Panel A: The central horizontal line of the box plot represents the median aPHQ-9 score, while the upper and lower edges of the box represent the IQR and the error bars represent 95% of the data range. The dots represent individual scores which are jittered horizontally in proportion to the number of data points at each score. Lines connect baseline and endline scores, when both are present. The Hodges-Lehmann median of differences is presented alongside a continuity-corrected 95% CI and p value estimated using the Wilcoxon rank-sum test. Panel B: The bars represent the percentage of participants at each level of reduction in aPHQ-9 score between baseline and endline.

**Table 2.** Intervention effects on primary and secondary outcomes ^1^.

|  | CBT | Control | Comparison<sup>2</sup><br>(95% CI) | P value<sup>3</sup> |
| --- | --- | --- | --- | --- |
| Primary outcome | n = 75 | n = 65 |  |  |
| Endline aPHQ-9 | 2 (1, 4) | 7 (3, 9) | -4 (-5, -3) | <0.001 |
| Secondary outcomes | n = 75 | n = 65 |  |  |
| Change in aPHQ-9, endline - baseline | -9 (-11, -7) | -4 (-7, -2) | -4 (-6, -3) | <0.001 |
| Reduction in aPHQ-9 from baseline, n (%) |  |  |  |  |
| ≤ 3 | 3 (4) | 29 (45) | 0.05 (0.01, 0.16) | <0.001 |
| > 3 | 72 (96) | 36 (55) | 19.33 (6.33, 84.59) | <0.001 |
| > 5 | 66 (88) | 26 (40) | 11.00 (4.86, 27.22) | <0.001 |
| > 9 | 30 (40) | 4 (6) | 10.17 (3.69, 36.05) | <0.001 |
| > 50% | 68 (91) | 29 (45) | 12.06 (5.06, 32.48) | <0.001 |
| Endline aPHQ-9 < 5 | 59 (79) | 22 (34) | 7.21 (3.46, 15.71) | <0.001 |
| Postnatal outcomes among antenatal enrollees by visit |  |  |  |  |
| 1.5 mo postnatal | n = 60 | n = 53 |  |  |
| aPHQ-9 score | 2 (0, 3) | 3 (0, 6) | -2 (-3, 0) | 0.007 |
| aPHQ-9 < 5, n (%) | 49 (82) | 29 (55) | 3.69 (1.61, 8.88) | 0.003 |
| 3 mo postnatal | n = 59 | n = 49 |  |  |
| aPHQ-9 score | 2 (0, 4) | 4 (1, 7) | -1 (-3, 0) | 0.010 |
| aPHQ-9 < 5, n (%) | 49 (83) | 26 (53) | 4.33 (1.84, 10.85) | 0.001 |
| 6 mo postnatal | n = 53 | n = 44 |  |  |
| aPHQ-9 score | 2 (0, 4) | 3 (0, 6) | -1 (-2, 0) | 0.18 |
| aPHQ-9 < 5, n (%) | 40 (76) | 27 (61) | 1.94 (0.81, 4.70) | 0.14 |
| 9 mo postnatal | n = 48 | n = 36 |  |  |
| aPHQ-9 score | 0 (0, 3) | 2 (1, 5) | -1 (-2, 0) | 0.006 |
| aPHQ-9 < 5, n (%) | 43 (90) | 26 (72) | 3.31 (1.05, 11.63) | 0.047 |

<sup>1</sup> Values are median (IQR) unless otherwise indicated.
<sup>2</sup> For continuous outcomes, this represents the Hodges-Lehman median of differences, with 95% confidence intervals calculated using normal approximation with continuity correction. Values < 0 indicate lower scores for CBT compared with control. For binary outcomes, this represents an odds ratio, estimated using logistic regression. Values > 1 indicate greater odds for CBT relative to control.
<sup>3</sup> For continuous outcomes, p values were estimated using the Wilcoxon rank-sum test. For binary outcomes, the Wald test was used to estimate p values.

Similarly, the proportion of CBT participants who achieved remission was higher than in the control group at 1.5, 3, and 9 months postpartum, but not at 6 months. In two post-hoc sensitivity analyses of imputed endline aPHQ-9 data for those without endline aPHQ-9 scores, results were similar to the primary mITT analysis (Appendix 3 and 4).

In a prespecified secondary analysis, adjustment for maternal age, education level, and baseline aPHQ-9 score did not meaningfully alter findings of unadjusted analyses (Appendix 5). In the prespecified subgroup analysis, we compared endline aPHQ-9 scores of participants by age at enrollment, reported education level, baseline aPHQ-9, and nutritional supplement randomized group in COGENT and found no statistically significant effect (Figure 3).

**Figure 3.**
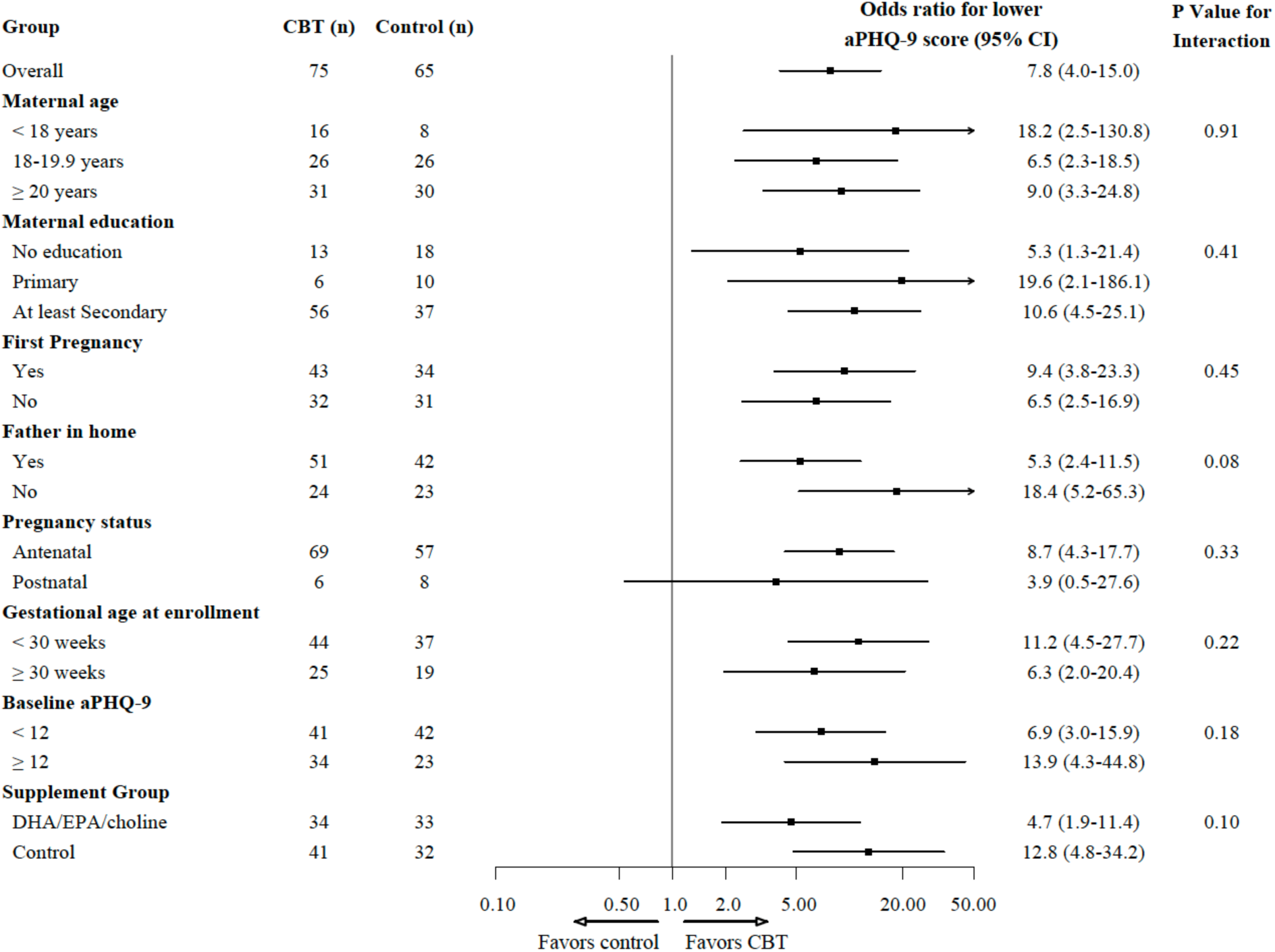
Forest plot. Odds ratio of a higher adapted Patient Health Questionnaire-9 score at endline in the CBT group compared with the control group (modified intention-to-treat population) by selected sub-groups. Odds ratios are represented by black boxes and 95% CIs by lines extending horizontally to each side of the black box. These were estimated using ordinal logistic regression and are displayed overall and for each sub-group. P values are shown for the interaction term between intervention group and subgroup variable. For maternal age and baseline aPHQ-9 score, the pre-specified analysis plan was for modeling these variables as continuous; the groupings shown are post-hoc. CBT=cognitive behavioral therapy. DHA=docosahexaenoic acid. EPA=eicosapentaenoic acid.

In a post-hoc analysis, additional subgroups first pregnancy vs. other, presence of father in home, and gestational age at CBT enrollment among participants enrolled antenatally were also analyzed. No statistically significant effect heterogeneity was identified.

Inter-assessor reliability was assessed at two times during the trial. At the first timepoint, 71 participants underwent repeat aPHQ-9 testing and the ICC was 0.94 (95% CI 0.91, 0.97). At the second timepoint, 51 participants under repeat aPHQ-9 testing and the ICC was 0.91 (95% CI 0.84, 0.97).

## DISCUSSION

In this randomized controlled trial conducted in rural Sierra Leone, a culturally adapted CBT intervention delivered by trained community counselors significantly reduced depressive symptoms among pregnant and postpartum women compared to standard care. The 4-point median reduction in aPHQ-9 scores represents a clinically meaningful improvement, with 79% of CBT participants achieving remission compared to 34% in the control group. Notably, these benefits were sustained through nine months postpartum, demonstrating lasting effects of the intervention.

The effectiveness of CBT for perinatal depression has been well-established, and our findings align with this evidence base while extending it to a novel population and setting. The magnitude of our effect represents a clinically meaningful improvement, comparable to effects observed in other CBT trials for perinatal depression.^16^ However, our study advances the field by demonstrating effectiveness in a population with very low literacy rates, where standard CBT protocols requiring written materials would be unfeasible.

Our findings contribute to the growing evidence for community-delivered mental health interventions in low-resource settings. Similar to the Thinking Healthy Programme in India, where peer women delivered CBT-based interventions with significant improvements in perinatal depression remission at six months postpartum^5^, our study demonstrates that local community members can effectively deliver psychological interventions. The Friendship Bench intervention in Zimbabwe similarly showed that lay health workers could achieve substantial symptom improvements for common mental disorders at six-month follow-up.^6^ Our study extends these findings by specifically addressing the challenge of adaptation for rural populations and accommodation for non-literate participants. The intervention was culturally adapted to align with local beliefs and social structures while literacy barriers were addressed by replacing written components with oral processing, role-play, and pictorial methods while maintaining core CBT principles.^8^

The sustained effects observed at nine months postpartum distinguish our findings from many intervention studies that show diminishing benefits over time. Prior studies of CBT for perinatal depression in high-income settings have demonstrated mixed results regarding long-term sustainability, with some maintaining significant effects at 6-12 month follow up^17,18^, while others have found diminishing effects over time, particularly when interventions were brief or lacked adequate skills consolidation^19^. In contrast to previous studies in sub-Saharan Africa that primarily examined short-term outcomes immediately post-intervention with limited evidence of longer-term follow-up^20^, our study is among the first to demonstrate sustained benefits of culturally adapted CBT at nine months follow-up in a rural African population. Unlike prior adaptations that focused primarily on language translation and surface-level modifications^21^, our adaptation specifically addressed literary barriers by including oral delivery methods, pictorial representations, integration with local conceptual frameworks, and accommodation of traditional value systems which may have enhanced skill retention and long-term effectiveness. This durability suggests that the skills taught in our adapted CBT program were successfully internalized and continued to benefit participants long after treatment completion, which has important implications for cost-effectiveness and public health impact.

In addition to the direct benefits for maternal mental health, our findings likely have important implications for early childhood outcomes. Poor maternal mental health during pregnancy and after birth is associated with adverse birth outcomes, impaired infant growth, and delays in cognitive, motor, language, socio-emotional development.^22–26^ Maternal mental health interventions—including CBT, psychosocial support, and community health worker-delivered programs—have been shown in low- and middle-income settings to improve children’s developmental and socio-emotional outcomes, increase exclusive breastfeeding, and, in some studies, reduce risk of undernutrition or growth faltering.^27–30^ These benefits are especially relevant in settings like Pujehun, Sierra Leone, where rates of childhood malnutrition and limited access to early learning opportunities remain high. In such contexts, early developmental delays often lead to persistent learning difficulties, reduced educational attainment, and long-term health consequences.^31–34^ The effectiveness of our intervention suggests potential not only for improving maternal well-being, but also for promoting intergenerational gains in child development and growth.

While this study demonstrates that CBT can be successfully adapted for a rural, largely non-reading population, scalability will require high levels of community engagement and adequate resources for effective delivery. Although participants agreed to return to the clinic for their CBT sessions, attendance was often inconsistent. The frequent follow-up visits likely contributed to the intervention’s success, though this comes with an economic cost for participants, many of whom are subsistence farmers. Home visits by the study team were necessary to complete many of the sessions, which would not be feasible without full-time staff having reliable transportation to rural communities. This highlights the need for dedicated, trained personnel and flexible delivery models to ensure sustainability at scale.

This study also highlighted the potential of task-shifting mental health interventions to women in the communities they serve. In our study, local women were selected and trained to deliver CBT, showing that lay providers can effectively support maternal mental health interventions. Community health workers and other lay counselors could be engaged in future efforts to expand this model. This aligns with successful task-shifting models such as the Thinking Healthy Programme, where community health workers delivered CBT-based interventions for perinatal depression in Pakistan and India, and the Friendship Bench intervention in Zimbabwe, where trained lay health workers provided psychological support for common mental disorders. There is an opportunity to build on existing structures, such as women’s groups or peer support networks, to strengthen reach and engagement. Successful scaling of this model will rely on dedicated funding and integration into existing community and health systems. Future research should explore large-scale implementation strategies.

This study’s randomized controlled design strengthens the validity of our findings. The trial was implemented in a rural Sub-Saharan African setting with limited access to mental health care and low literacy levels, illustrating the feasibility of adapting psychological treatments for illiterate, low-resource communities. We achieved high follow-up rates, strengthening our results, and the intervention was delivered by local women, trained and supervised by the Mental Health Coalition in Sierra Leone, enhancing CBT fidelity. Another strength of this study was the cultural adaptation of the CBT model, tailored to fit the educational and local context of the population, which contributed to its high acceptability and engagement.

Our study has several limitations. Despite measures used to facilitate blinding of outcome assessors, full masking may have been compromised. Participants receiving CBT attended weekly sessions, while those in the control arm were only seen during their assigned clinic visits for the full COGENT study–every two weeks during pregnancy and more spaced apart during the postpartum period. This difference in visit frequency may have revealed group allocation to outcome assessors who were present at each clinic visit. Additionally, home visits were necessary due to missed sessions, and counselors often traveled together to the participant’s village. Only the counselor assigned to conduct CBT delivered the session, but the proximity could have increased the risk of unblinding the outcome assessor. Another limitation of these findings is that literacy assessment was not conducted at study onset, but occurred later which results in some missing data and limiting our ability to fully characterize the literacy levels of all participants at baseline. Finally, the generalizability of our findings is limited and results may not translate to urban settings or other cultural contexts without further adaption.

In conclusion, we found that a culturally adapted novel CBT program was an effective intervention for women with peripartum depression. While our findings demonstrate an adapted CBT’s success and scalability potential, further research is needed to understand and address potential implementation constraints in other settings.

## Supporting information

Protocol & Tables

## Data Availability

All data produced in the present study are available upon reasonable request to the authors

