## Supplementary material for "A cognitive behavioral therapy program designed for low-literacy end-users with perinatal depression in Sierra Leone: an investigator- and outcomes assessor-blinded, controlled randomized trial": Protocol & Tables

### COGENT - Improving COgnition and GEstational duration with targeted NuTrition

**National Clinical Trial (NCT) Identified Number: TBD**

**Principal Investigator: Mark Manary, MD**

**Washington University in St. Louis**

**Funded by: Open Philanthropy**

**Version Number: 1**

**21 April 2023**

#### Summary of Changes from Previous Version:

| Affected Section(s) | Summary of Revisions Made | Rationale |
|---|---|---|

#### Table of Contents

STATEMENT OF COMPLIANCE ... 1
1 PROTOCOL SUMMARY ... 2
1.1 Synopsis ... 2
1.2 Schema ... 3
1.3 Schedule of Activities (SoA) ... 5
2 INTRODUCTION ... 7
2.1 Study Rationale ... 7
2.2 Background ... 7
2.3 Risk/Benefit Assessment ... 9
2.3.1 Known Potential Risks ... 9
2.3.2 Known Potential Benefits ... 10
2.3.3 Assessment of Potential Risks and Benefits ... 11
3 OBJECTIVES AND ENDPOINTS ... 12
4 STUDY DESIGN ... 15
4.1 Overall Design ... 15
4.2 Scientific Rationale for Study Design ... 15
4.3 Justification for Dose ... 16
4.4 End of Study Definition ... 16
5 STUDY POPULATION ... 17
5.1 Inclusion Criteria ... 17
5.2 Exclusion Criteria ... 17
5.3 Lifestyle Considerations ... 18
5.4 Screen Failures ... 18
5.5 Strategies for Recruitment and Retention ... 18
6 STUDY INTERVENTION ... 20
6.1 Study Intervention(s) Administration ... 20
6.1.1 Study Intervention Description ... 20
6.1.2 Dosing and Administration ... 22
6.2 Preparation/Handling/Storage/Accountability ... 22
6.2.1 Acquisition and accountability ... 22
6.2.2 Formulation, Appearance, Packaging, and Labeling ... 23
6.2.3 Product Storage and Stability ... 23
6.2.4 Preparation ... 24
6.3 Measures to Minimize Bias: Randomization and Blinding ... 24
6.4 Study Intervention Compliance ... 25
6.5 Concomitant Therapy ... 26
6.5.1 Rescue Medicine ... 26
7 STUDY INTERVENTION DISCONTINUATION AND PARTICIPANT DISCONTINUATION/WITHDRAWAL ... 27
7.1 Discontinuation of Study Intervention ... 27
7.2 Participant Discontinuation/Withdrawal from the Study ... 27
7.3 Lost to Follow-Up ... 28
8 STUDY ASSESSMENTS AND PROCEDURES ... 29
8.1 Efficacy Assessments ... 29
8.2 Safety and Other Assessments ... 32
8.3 Adverse Events and Serious Adverse Events ... 33

8.3.1 Definition of Adverse Events (AE) ... 33
8.3.2 Definition of Serious Adverse Events (SAE) ... 33
8.3.3 Classification of an Adverse Event ... 33
8.3.4 Time Period and Frequency for Event Assessment and Follow-Up ... 35
8.3.5 Adverse Event Reporting ... 35
8.3.6 Serious Adverse Event Reporting ... 36
8.3.7 Reporting Events to Participants ... 37
8.3.8 Events of Special Interest ... 37
8.3.9 Reporting of Pregnancy ... 37
8.4 Unanticipated Problems ... 37
8.4.1 Definition of Unanticipated Problems (UP) ... 37
8.4.2 Unanticipated Problem Reporting ... 37
8.4.3 Reporting Unanticipated Problems to Participants ... 38
9 STATISTICAL CONSIDERATIONS ... 39
9.1 Statistical Hypotheses ... 39
9.2 Sample Size Determination ... 39
9.3 Populations for Analyses ... 44
9.4 Statistical Analyses ... 44
9.4.1 General Approach ... 44
9.4.2 Analysis of the Primary Efficacy Endpoint(s) ... 45
9.4.3 Analysis of the Secondary Endpoint(s) ... 45
9.4.4 Safety Analyses ... 47
9.4.5 Baseline Descriptive Statistics ... 47
9.4.6 Planned Interim Analyses ... 47
9.4.7 Sub-Group Analyses ... 47
9.4.8 Tabulation of Individual participant Data ... 48
9.4.9 Exploratory Analyses ... 48
10 SUPPORTING DOCUMENTATION AND OPERATIONAL CONSIDERATIONS ... 48
10.1 Regulatory, Ethical, and Study Oversight Considerations ... 48
10.1.1 Informed Consent Process ... 48
10.1.2 Study Discontinuation and Closure ... 49
10.1.3 Confidentiality and Privacy ... 49
10.1.4 Future Use of Stored Specimens and Data ... 50
10.1.5 Key Roles, Study team, and Study Governance ... 51
10.1.6 Safety Oversight ... 51
10.1.7 Clinical Monitoring ... 51
10.1.8 Quality Assurance and Quality Control ... 52
10.1.9 Data Handling and Record Keeping ... 52
10.1.10 Protocol Deviations ... 53
10.1.11 Publication and Data Sharing Policy ... 53
10.1.12 Conflict of Interest Policy ... 53
10.2 Additional Considerations ... 53
10.3 Abbreviations ... 54
10.4 Protocol Amendment History ... 55
11 REFERENCES ... 56

#### STATEMENT OF COMPLIANCE

The trial will be conducted in accordance with International Conference on Harmonisation Good Clinical Practice (ICH GCP) and applicable United States (US) Code of Federal Regulations (CFR). The Principal Investigator will assure that no deviation from or changes to the protocol will take place without prior agreement and documented approval from the Institutional Review Board (IRB), except where necessary to eliminate an immediate hazard to the trial participants.

The protocol, informed consent forms, recruitment materials, and all participant materials will be submitted to the IRB for review and approval. Approval of both the protocol and the consent form must be obtained before any participant is enrolled. Any amendment to the protocol will require review and approval by the IRB before the changes are implemented to the study. All changes to the consent form will be IRB approved; a determination will be made regarding whether a new consent needs to be obtained from participants who provided consent, using a previously approved consent form.

### 1 PROTOCOL SUMMARY

#### 1.1 SYNOPSIS

| | |
|---|---|
| **Title:** | COGENT - Improving COgnition and GEstational duration with targeted NuTrition |
| **Study Description:** | A randomized, investigator-blinded, controlled clinical trial of factorial design among malnourished pregnant women in Sierra Leone testing the hypotheses that (1) adding 500mg docosahexaenoic acid (DHA), 500mg eicosapentaenoic acid (EPA), and 550mg choline to a supplementary food (maternal ready-to-use supplementary food +, M-RUSF+) will prolong gestation and improve infant cognitive development compared with a similar supplementary food but without DHA/EPA/choline (M-RUSF), and (2) a novel cognitive behavioral therapy (CBT) will improve depression among women with ante- or post-natal depression compared with no CBT. |
| **Objectives:** | Primary Objectives. Comparison 1: To determine whether M-RUSF+ provided to malnourished pregnant Sierra Leonean women will prolong gestation and improve infant cognitive development at 9 months of age when compared with provision of M-RUSF. Comparison 2: To determine whether provision of a novel CBT designed for illiterate end-users will improve depressive symptoms among malnourished pregnant women with ante- or post-natal depression compared with no CBT.<br>Secondary Objectives: To determine whether M-RUSF+ will reduce early and late pre-term birth, improve birth length/weight, reduce low birth weight, improve sub-domain cognitive development scores, reduce neonatal mortality, and reduce depression scores compared with M-RUSF. |
| **Endpoints:** | Primary Endpoints: Comparison 1: Gestational duration (days), Malawi Developmental Assessment Tool (MDAT) global z-score. Comparison 2: Patient Health Questionnaire-9 score.<br>Secondary Endpoints: Early pre-term (<34 weeks) and late pre-term (<37 weeks) birth, birth weight and length, low birth weight, neonatal mortality, PHQ-9 scores among all participants, MDAT sub-domain scores. |
| **Study Population:** | 1600 pregnant persons ≥ 13 years of age with mid-upper arm circumference ≤ 23.0 cm or body-mass index < 18.5 who seek care at a participating antenatal clinic in Bo and Pujehun Districts in Southern Sierra Leone. Gestational duration will be compared among participants with enrollment GA ≤ 30 weeks who have singleton live births. |
| **Description of Sites Enrolling Participants:** | The study will take place at 30-40 antenatal clinics operated by the Sierra Leonean government. |
| **Description of Study Intervention:** | The intervention and control supplementary foods will be provided in 100g sachets. They will be composed of peanuts, sugar, palm oil, non-fat milk powder, and pearl millet, for 520 Kcal/day, 18g protein, 33g fat. The intervention food will also contain 500mg DHA, 500mg EPA, and 550mg choline. CBT will be performed weekly for 6 sessions. |
| **Study Duration:** | 30 months |
| **Participant Duration:** | 10-16 months |

#### 1.2 SCHEMA

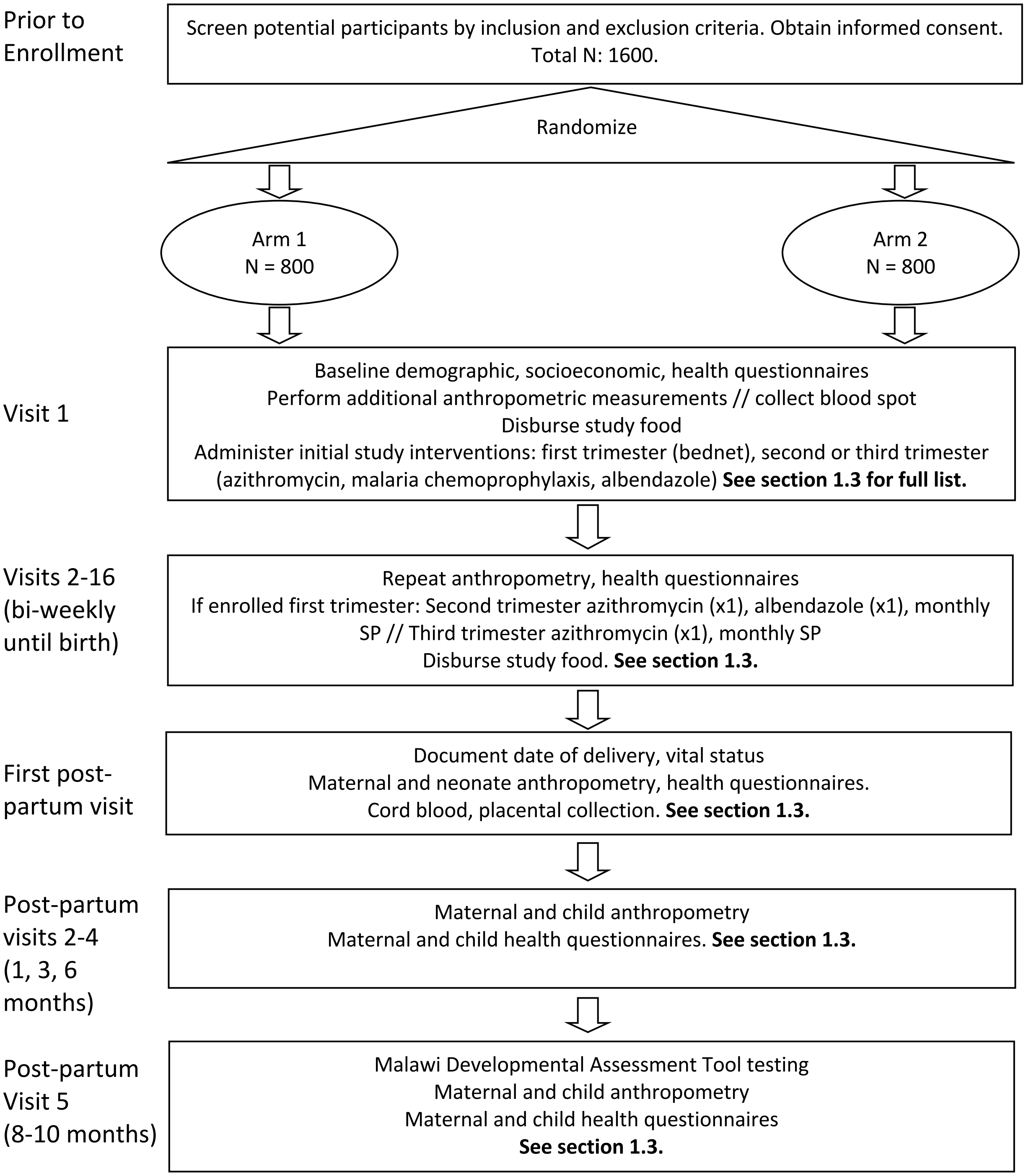
Prior to Enrollment
Screen potential participants by inclusion and exclusion criteria. Obtain informed consent. Total N: 1600.
Randomize
Arm 1
N = 800
Arm 2
N = 800
Visit 1
Baseline demographic, socioeconomic, health questionnaires
Perform additional anthropometric measurements // collect blood spot
Disburse study food
Administer initial study interventions: first trimester (bednet), second or third trimester (azithromycin, malaria chemoprophylaxis, albendazole) See section 1.3 for full list.
Visits 2-16 (bi-weekly until birth)
Repeat anthropometry, health questionnaires
If enrolled first trimester: Second trimester azithromycin (x1), albendazole (x1), monthly SP // Third trimester azithromycin (x1), monthly SP
Disburse study food. See section 1.3.
First post-partum visit
Document date of delivery, vital status
Maternal and neonate anthropometry, health questionnaires.
Cord blood, placental collection. See section 1.3.
Post-partum visits 2-4 (1, 3, 6 months)
Maternal and child anthropometry
Maternal and child health questionnaires. See section 1.3.
Post-partum Visit 5 (8-10 months)
Malawi Developmental Assessment Tool testing
Maternal and child anthropometry
Maternal and child health questionnaires
See section 1.3.

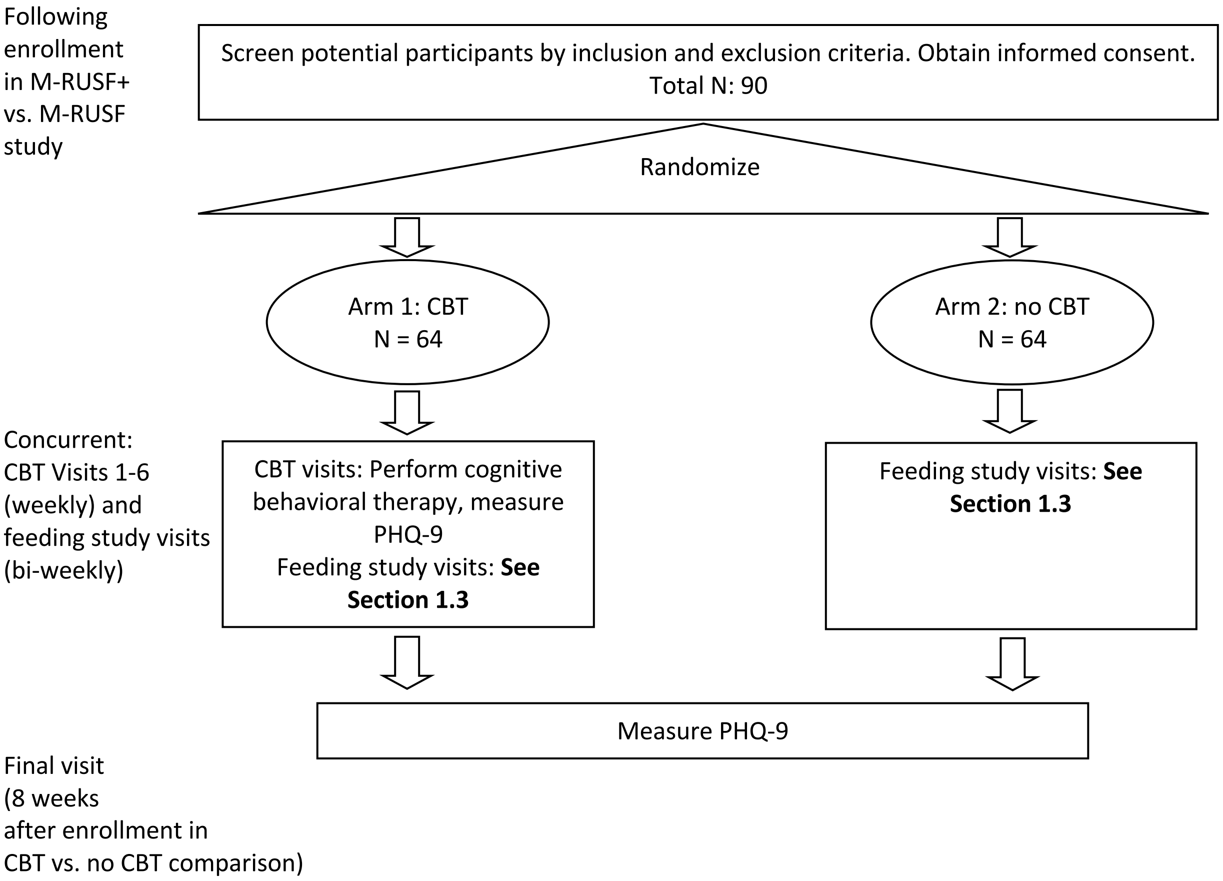
Following enrollment in M-RUSF+ vs. M-RUSF study
Screen potential participants by inclusion and exclusion criteria. Obtain informed consent.
Total N: 90
Randomize
Arm 1: CBT
N = 64
Arm 2: no CBT
N = 64
Concurrent:
CBT Visits 1-6 (weekly) and feeding study visits (bi-weekly)
CBT visits: Perform cognitive behavioral therapy, measure PHQ-9
Feeding study visits: See Section 1.3
Feeding study visits: See Section 1.3
Measure PHQ-9
Final visit (8 weeks after enrollment in CBT vs. no CBT comparison)

#### 1.3 SCHEDULE OF ACTIVITIES (SOA)

Trial of DHA/EPA/choline-containing supplementary food (M-RUSF+) vs. M-RUSF

<table>
<tr><th rowspan="3">Procedures</th><th colspan="2">Enrollment (≤ 30 weeks gestation)</th><th colspan="3">Bi-weekly prenatal visits</th><th rowspan="3">First post-natal visit</th><th rowspan="3">1mo, 3mo, 6mo post-natal visits</th><th rowspan="3">9mo post-natal visit (8 – 10 months)</th></tr>
<tr><th rowspan="2">1st trimester</th><th rowspan="2">2nd/3rd trimester</th><th rowspan="2">1st trimester</th><th colspan="2">2nd / 3rd trimester</th></tr>
<tr><th>First visit</th><th>Subsequent visits</th></tr>
<tr><td>Informed consent</td><td colspan="2">X</td><td></td><td></td><td></td><td></td><td></td><td></td></tr>
<tr><td>Demographic questionnaire</td><td colspan="2">X</td><td></td><td></td><td></td><td></td><td></td><td></td></tr>
<tr><td>Socioeconomic questionnaire</td><td colspan="2"></td><td></td><td></td><td></td><td></td><td></td><td></td></tr>
<tr><td>Medical history</td><td colspan="2">X</td><td></td><td></td><td></td><td></td><td></td><td></td></tr>
<tr><td>Randomization</td><td colspan="2">X</td><td></td><td></td><td></td><td></td><td></td><td></td></tr>
<tr><td>Ultrasound gestational age</td><td colspan="2">X</td><td></td><td></td><td></td><td></td><td></td><td></td></tr>
<tr><td>Disburse study food</td><td colspan="2">X</td><td>X</td><td>X</td><td>X</td><td></td><td></td><td></td></tr>
<tr><td>Disburse bednets</td><td colspan="2">X</td><td></td><td></td><td></td><td></td><td></td><td></td></tr>
<tr><td>Azithromycin administration[1]</td><td></td><td>X</td><td></td><td>X</td><td></td><td></td><td></td><td></td></tr>
<tr><td>Albendazole administration[1]</td><td></td><td>X</td><td></td><td>X</td><td></td><td></td><td></td><td></td></tr>
<tr><td>Sulfadoxine / pyrimethamine[1]</td><td></td><td>X</td><td></td><td>X</td><td>X (monthly)</td><td></td><td></td><td></td></tr>
<tr><td>Maternal anthropometry</td><td colspan="2">X</td><td>X</td><td>X</td><td>X</td><td>X</td><td>X</td><td>X</td></tr>
<tr><td>Maternal symptom questionnaire</td><td colspan="2">X</td><td>X</td><td>X</td><td>X</td><td>X</td><td>X</td><td>X</td></tr>
<tr><td>Food insecurity questionnaire</td><td colspan="2">X</td><td></td><td></td><td></td><td></td><td></td><td></td></tr>
<tr><td>Dietary survey</td><td colspan="2">X</td><td></td><td></td><td></td><td></td><td></td><td></td></tr>
<tr><td>Blood pressure</td><td colspan="2">X</td><td>X</td><td>X</td><td>X</td><td></td><td></td><td></td></tr>
<tr><td>PHQ-9</td><td colspan="2">X</td><td>X</td><td>X</td><td>X</td><td>X</td><td>X</td><td></td></tr>
<tr><td>Blood spot collection[2]</td><td colspan="2">X</td><td></td><td></td><td>X (once)</td><td></td><td></td><td></td></tr>
<tr><td>Cord blood collection[3]</td><td colspan="2"></td><td></td><td></td><td></td><td>X</td><td></td><td></td></tr>
<tr><td>Placental sample collection</td><td colspan="2"></td><td></td><td></td><td></td><td>X</td><td></td><td></td></tr>
<tr><td>Placental weight measurement</td><td colspan="2"></td><td></td><td></td><td></td><td>X</td><td></td><td></td></tr>
<tr><td>Child anthropometry</td><td colspan="2"></td><td></td><td></td><td></td><td>X</td><td>X</td><td>X</td></tr>
<tr><td>Child symptom questionnaire</td><td colspan="2"></td><td></td><td></td><td></td><td>X</td><td>X</td><td>X</td></tr>
<tr><td>MDAT</td><td colspan="2"></td><td></td><td></td><td></td><td></td><td></td><td>X</td></tr>
<tr><td>Adverse event review and evaluation</td><td colspan="2">X</td><td>X</td><td>X</td><td>X</td><td>X</td><td>X</td><td>X</td></tr>
</table>

[1] Albendazole to be given once, either upon enrollment if enrolled during 2nd / 3rd trimester, or at first 2nd trimester visit if enrolled during 1st trimester. Azithromycin to be given 2 times, once during 2nd trimester and once during third trimester. SP to be given monthly starting in 2nd trimester.

[2] A second blood spot will be collected within the 32-36-week gestational age

[3] Cord blood and placental samples will be collected when possible from mothers at hospitals/clinics with proximity to allow for research staff to be present for collection.

Trial of CBT vs. no CBT for ante- or postnatal depression.

Antenatal Depression. Intervention Group receives CBT. Control = Control Group.

| Procedures | Diagnosis visit: PHQ-9 ≥ 9 Week 0 | Week 1 | Week 2 | Week 3 | Week 4 | Week 5 | Week 6 | Post-treatment assessment Week 8 |
|---|---|---|---|---|---|---|---|---|
| Mental Health Screening (PHQ-9) | Intervention and Control | Intervention | Intervention and Control | Intervention | Intervention and Control | Intervention | Intervention and Control | Intervention and Control |
| Functioning Questionnaire- | Intervention and Control | Intervention | Intervention | Intervention | Intervention | Intervention | Intervention | |
| Cognitive Behavioral Therapy | | Intervention | Intervention | Intervention | Intervention | Intervention | Intervention | |
| Adverse event review and evaluation | Intervention and Control | Intervention | Intervention And Control | Intervention | Intervention And Control | Intervention | Intervention And Control | Intervention and Control |

During the antenatal period, all participants are seen at clinic bi-weekly, during which time they will undergo PHQ-9 assessments. If they are diagnosed with depression, this becomes week 0 in the CBT vs. no CBT study.

Postnatal depression. Intervention Group receives CBT. Control = Control Group.

| Procedures | Diagnosis visit: PHQ-9 ≥ 9 Week 0 | Weeks 1-6 | Post-treatment assessment Week 8 |
|---|---|---|---|
| Mental Health Screening (PHQ-9) | Intervention and Control | Intervention | Intervention and Control |
| Functioning Questionnaire- | Intervention and Control | Intervention | |
| Cognitive Behavioral Therapy | | Intervention | |
| Adverse event review and evaluation | Intervention and Control | Intervention | Intervention and Control |

Postnatally, all participants are scheduled for study visits at birth and 6, 12, 24, and 36 weeks after birth. They will undergo PHQ-9 assessment at each scheduled visit, which may trigger enrollment in the CBT vs. no CBT study, and may take place when a participant is already enrolled in the CBT vs. no CBT study. Participants will be encouraged to come to clinic if they feel they are developing signs or symptoms of depression, whether on their preset schedule or not.

### 2 INTRODUCTION

#### 2.1 STUDY RATIONALE

1. Preterm birth remains common worldwide, is the leading risk factor for neonatal mortality, and is associated with short- and long-term developmental complications among those who survive. Many of these deaths occur in Sub-Saharan Africa (SSA); in Sierra Leone, the neonatal mortality rate is 31/1,000 live births.[1] The omega-3 long-chain polyunsaturated fatty acid (PUFA) docosahexaenoic acid (DHA) shows promise in reducing prematurity, but effects have been inconsistent. Choline is a 1-carbon metabolite that plays key roles in DHA trafficking and tissue integration. Maternal undernutrition, which impacts nearly one-quarter of pregnant women in Sub-Saharan Africa, is a state of total-body nutritional depletion that carries increased risks for adverse maternal and fetal outcomes, including preterm birth.[2,3] We hypothesize that adding omega-3 PUFA (DHA+EPA) and choline to a supplementary food in this high-risk population of malnourished pregnant women in rural Sierra Leone will prolong gestation and thereby reduce the most potent risk factor for neonatal mortality and morbidity. A positive finding would have implications for millions of women worldwide, for whom care guidelines are scant.

2. Maternal undernutrition is associated with offspring malnutrition, stunting, and impaired cognitive development. A randomized, controlled, blinded trial published by the PI's research group in 2021 demonstrated that child cognitive development is sensitive to the composition of fatty acid intake during episodes of undernutrition; specifically, cognitive improved when therapeutic food was formulated with lower omega-6 levels and added DHA.[4] We hypothesize adding omega-3 PUFA (DHA+EPA) and choline to a supplementary food provided to malnourished pregnant women in rural Sierra Leone will improve offspring cognitive development at 9 months of age as assessed by the Malawi Developmental Assessment Tool.

3. Ante- and postnatal depression are common yet underrecognized and undertreated in rural SSA, including in Sierra Leone. Cognitive behavioral therapy (CBT) is an evidence-based intervention for perinatal depression, yet current formulations cannot be applied to illiterate populations. In partnership with the Mental Health Coalition of Sierra Leone, the PI developed a novel CBT for use in illiterate populations. We hypothesize that this novel CBT will improve PHQ-9 scores among women diagnosed with ante- or postnatal depression during the aforementioned clinical trial. This will be a factorial trial.

#### 2.2 BACKGROUND

**Undernutrition and inflammation in pregnancy**

Nearly one-quarter of pregnant women in SSA are malnourished.[2] This is a high-risk state for them and their offspring and requires treatment. Quality supplemental nutrition is an essential but insufficient part of treatment in contexts of high inflammation, such as are common in rural SSA.[5] A recent large, randomized controlled clinical effectiveness trial in Sierra Leone led by the PI of this protocol/trial, hereafter referred to as MamaDutasi, demonstrated a 55% reduction in neonatal mortality as well as longer, heavier newborns when two doses of azithromycin and monthly malaria chemoprophylaxis were added to high-quality supplementary food during pregnancy.[6] Combining this anti-infective treatment bundle with further improvements in the supplementary food product has the potential to yield even greater benefits for these mothers and their children, as does treatment of ante- and postnatal depression.

**Docosahexaenoic acid and choline supplementation in pregnancy**

There is a large body of randomized controlled trial (RCT) evidence supporting DHA supplementation in pregnancy, particularly as regards prevention of early preterm birth and especially favoring higher-dose supplementation (>600mg/day) in those with low DHA status at baseline.[7] A 2018 Cochrane review and a 2021 update found that omega-3 PUFA supplementation during pregnancy is likely to reduce preterm birth (analysis included 27,726 participants) and early preterm birth (16,782 participants), prolong gestation (23,359 participants), increase birth weight (20,104 participants), and reduce serious adverse events for neonate/infant, including perinatal death.[8] Findings have not been entirely consistent, however; for example, a large RCT in Australia did not demonstrate a reduction in early preterm birth with omega-3 supplementation, though rates were low and DHA status at baseline was higher than has been seen in other studies.[9,10] The DHA status of malnourished pregnant women in Sierra Leone is unknown. While people in some areas of Sierra Leone may have consistent fish consumption, which would be expected to raise DHA levels depending on type of fish, others do not. It is plausible that DHA status will be low in the setting of undernutrition and pregnancy. Thus, it is possible that supplementation with omega-3 in this likely deficient state might prolong gestation. As gestational duration is tightly linked with neonatal mortality and subsequent development issues, prolonging gestation is a possible path for benefit in this population. The following figure shows this relationship as seen in MamaDutasi (figure unpublished).

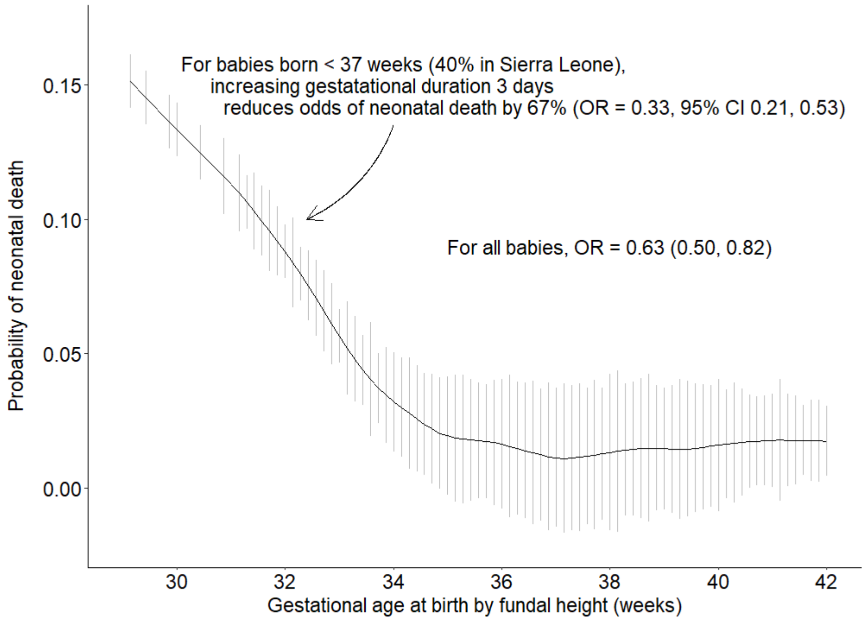

The evidence is more mixed for the effects of prenatal omega-3 supplementation on infant cognitive development. Decades of findings from epidemiological studies and laboratory science, including with animal models, support the essential role of DHA in the structure and function of the brain and retina.[11] Half of the brain's DHA is accumulated during gestation, a process with rapidly increases in the third trimester.[12-14] DHA ultimately composes 10-20% of the brain's lipids by weight. Breast milk contains large amounts of DHA, suggesting its importance for early brain development. In contrast to these lines of evidence, the 2018 Cochrane review did not find benefit for DHA supplementation in pregnancy across a variety of child cognitive measures.

Two recent studies suggest DHA supplementation's benefits may be context-dependent. An RCT of very pre-term infants demonstrated improved general intelligence 5 years after DHA supplementation during infancy.[15] While supplementation was not prenatal in this study, the results do suggest the importance of DHA during brain development early in life, particularly in the third trimester. The Improved PUFA study, completed by our research group in 2021, offers another such example of cognitive development's sensitivity to fatty acid intake. In the study, children with severe acute malnutrition who received ready-to-use therapeutic food (RUTF) with reduced omega-6 and greater omega-3, including DHA and eicosapentaenoic acid (EPA), demonstrated improved cognitive development compared with a standard RUTF without added omega-3s six months after treatment.[4] Again, while this was not a study of prenatal DHA supplementation, it does support a role for DHA in the setting of undernutrition and likely deficiency. It is possible that DHA transfer to the fetus is inadequate in the setting of maternal undernutrition. Given the rapidity and amount of DHA accumulation in the brain during gestation, this window might present an opportunity for supplementation.

**Choline, DHA, and pregnancy**

Choline is essential for a variety of physiologic processes, including trafficking and incorporation of DHA into cell membranes. Choline deficiency induces a host of developmental problems in animal studies and has been shown in several small RCTs to improve measures of cognition in children. A recent RCT showed that prenatal choline supplementation improves hepatic DHA export and multiple markers of DHA status in pregnant women.[16] It is possible that co-supplementation with choline and DHA might magnify the benefits of both when given to a population at high risk for deficiencies.

**Cognitive behavioral therapy for ante- and postnatal depression**

The prevalence of antenatal depression has been found to be approximately 7-20%, while that of postpartum depression is nearly 20%. While prevalence assessments are scarce in Sub-Saharan Africa and non-existent in Sierra Leone, evidence suggests rates are higher in low- and middle-income countries. Both pre- and postnatal depression are associated with failure to seek care, poor diet, use of harmful substances, self-harm and attempted suicide, poor care of offspring and adverse effects on fetal to adolescent development. A 2020 systemic review of 5 RCTs found evidence supporting CBT for antenatal depression.[17] A 2016 systemic review found evidence for increased likelihood of remission with CBT among women with ante- and postnatal depression compared with usual care.[18] A 2018 systematic review of 20 RCTs found that CBT improved postpartum depression.[19] In all cases, CBT was applied in literate populations. Whether a novel CBT designed for use in the setting of illiteracy might benefit pregnant women with ante- or postnatal depression is unknown.

#### 2.3 RISK/BENEFIT ASSESSMENT

##### 2.3.1 KNOWN POTENTIAL RISKS

With respect to omega-3 PUFA/choline addition to a supplementary food, we do not anticipate any harms to participants. The amount of omega-3 PUFA to be given, 1g/day of DHA+EPA, is lower than what is contained in a 6-ounce serving of salmon. Similarly, the 550mg of choline to be included approximates 4 eggs' worth. In addition, the target population – malnourished pregnant women – is likely to be deficient in these nutrients, as they are malnourished and their diets contain few animal-source foods. Thus, any theoretical risks of over-supplementation are unlikely. In addition, omega-3 PUFAs have been studied as pregnancy supplements in RCTs including over 23,000 women. A 2018 Cochrane review found no evidence for harm in either mothers or offspring.[7] Below, several considerations are explicated in more detail.

Omega-3 PUFA might increase the incidence of pregnancies continuing beyond 42 weeks gestation, which itself increases risk for adverse birth outcomes. Among >16,000 RCT participants included in a 2021 meta-analysis, the relative risk for >42 weeks gestation with omega-3 supplementation vs. none was 1.31 (1.01, 1.70).[8] It is unclear, though, whether this potential risk would translate to malnourished pregnant women in Sierra Leone, a population different from those included in the studies which were meta-analyzed. Because of a lack of ultrasound capabilities, quality data on gestational duration in Sierra Leone are not available. In our recent RCT, fundal height was used to estimate gestational age (Hendrixson); only 9/1431 participants had final fundal heights estimated at > 42 weeks. Indeed, the issue motivating this clinical trial is the suspected high incidence of pre-term birth, suggested by 82% of participants in the above trial with final fundal height < 37cm and ~20% rate of LBW. Thus, we estimate any risk of adverse events related to prolonged gestation to be minimal.

A 1992 RCT identified a trend of greater blood loss during delivery with omega-3 PUFA supplementation.[20] Supplementation in this trial was at a higher dose, 2.7g/day, than will be given in COGENT. This safety concern has not been replicated in subsequent large RCTs, which have used doses closer COGENT's 1g/day.

Several trials that used fish-source omega-3s reported higher rates gastrointestinal symptoms in pregnant women, including burping, constipation, and diarrhea. These symptoms will be tracked among participants.

Several trials have followed participants and their offspring greater than 10 years after the trial. No longer term / delayed risks have been identified.

Choline intakes that are greater than an order of magnitude higher than the supplementation planned in this trial have been associated with hypotension, body odor, sweating, vomiting, and salivation.[21] Doses from involved studies ranged from 7.5 – 16 g daily. The tolerable upper intake level for adults has been set at 3.5 g daily. The dose used in this study will be 550mg/day, as justified in **section 4.3.**

With respect to CBT for pre- and post-natal depression, there is the potential for therapy to cause harm in participants. CBT involves uncovering, probing, and working to gain control over unproductive thought patterns and behaviors. This can cause distress by its nature, as it asks participants to confront negative thoughts, emotions, and physical responses to, for instance, anxiety and depression. A 2018 study of 100 CBT practitioners showed, in the therapists' perceptions, 27% of patients experience distress and 9% worsening of symptoms, with 21% of such effects being severe/very severe, though only 5% were persistent.[22] Of note, nearly half of patients referenced had undergone psychiatric hospitalization and were on medications for a psychiatric condition, suggesting more severe mental health issues than might be seen in this trial's population. However, it is possible that some participants may experience distress, and possibly temporary worsening of depression, as a result of receiving CBT.

##### 2.3.2 KNOWN POTENTIAL BENEFITS

As identified in a 2018 Cochrane review and 2021 update, omega-3 LCPUFA supplementation during pregnancy is likely to reduce preterm birth (27,726 participants) and early preterm birth (16,782 participants), prolong gestation (23,359 participants), increase birth weight (20,104 participants), and reduce serious adverse events for neonate/infant, including perinatal death.

We hypothesize that supplementation of DHA/EPA during pregnancy in malnourished women could impart cognitive developmental benefits in offspring. Decades of epidemiological and animal studies research supports the essential role of DHA in cognitive development and high rate of accretion in the brain during gestation. While previous human studies research has shown inconsistent effects of prenatal DHA supplementation, none of this research was done among malnourished pregnant women.

A 2021 Cochrane review found low certainty evidence of a small benefit of omega-3 PUFAs for depressive symptoms compared to placebo. The evidence was stronger for supplementation with more EPA. We hypothesize that 500mg/500mg DHA/EPA supplementation during pregnancy might reduce PHQ-9 scores and incidence of ante- and postnatal depression among women likely to be deficient in omega-3 PUFAs.

The 3 systematic reviews previously detailed demonstrate evidence for benefit of CBT on ante- and/or postnatal depression.[17-19]

##### 2.3.3 ASSESSMENT OF POTENTIAL RISKS AND BENEFITS

Everyday consumption of omega-3 PUFAs and choline in some populations in amounts greater than the proposed supplementation in COGENT argues strongly in favor of the safety of this intervention, as do several large RCTs documenting safety. In addition, the target population of malnourished pregnant women are likely to be deficient in both nutrients, further reducing the theoretical risk of over-supplementation. In contrast, the evidence in favor of omega-3 PUFAs for prolonging gestation, increasing birth weight, and reducing serious adverse neonatal events is strong. The potential benefits outweigh the marginal risks.

Regarding offspring cognitive development, there is no evidence of harm to offspring with prenatal supplementation. While the likelihood of benefit is less clear than in the case of prolonging gestation, were it found to be beneficial, this would have significant implications for millions of women suffering from undernutrition, and their children.

While CBT has been associated with distress in patients, 25 RCTs have documented efficacy in ante- and postnatal depression without revealing concerning safety signals. Left untreated, ante- and postnatal depression can have devastating effects on mom and offspring. If the novel CBT were found to be beneficial, it could have implications for millions of women with untreated ante- and postnatal depression who do not have therapeutic options available. The potential benefits outweigh the marginal risks.

### 3 OBJECTIVES AND ENDPOINTS

| OBJECTIVES | ENDPOINTS | JUSTIFICATION FOR ENDPOINTS |
|---|---|---|
| Primary | | |
| To determine whether adding 1g DHA/EPA + 550mg choline to a supplementary food provided to malnourished pregnant Sierra Leonean women ≤ 30 weeks GA at enrollment will prolong gestation. | Gestational duration, days | In settings of high rates of prematurity, prolonging gestation improves offspring outcomes. Based on recent RCT data, prolonging gestation by 3 days reduced odds of neonatal death by 67%. |
| To determine whether adding 1g DHA/EPA + 550mg choline to a supplementary food provided to malnourished pregnant Sierra Leonean women will improve offspring cognitive development. | Malawi Developmental Assessment Tool global z-score | By combining gross motor, fine motor, language, and social developmental assessments, MDAT provides a broad measure of child development. MDAT was developed in Malawi and has been validated in other Sub-Saharan African settings. |
| To determine whether provision of a novel cognitive behavioral therapy intervention will improve depressive symptoms among women with ante- or postnatal depression. | Patient Health Questionnaire-9 (PHQ-9) score | PHQ-9 has been validated as a screening and diagnostic tool for ante- and postnatal depression, including in Sub-Saharan Africa. It is easy to use, has been translated to many settings, and has demonstrated good sensitivity to changes in depression over time. PHQ-9 is a patient-centered outcome. |
| Secondary | | |
| To determine whether adding 1g DHA/EPA + 550mg choline to a supplementary food provided to malnourished pregnant Sierra Leonean women ≤ 30 weeks GA at enrollment will: | | |
| • Reduce early pre-term delivery. | Proportion of deliveries < 34 weeks gestation | ePTB carries a high risk of neonatal morbidity and mortality as well as neurodevelopmental sequelae. |
| • Improve birth weight and length | Birth weight (g), length (cm) | Birth weight predicts neonatal outcomes; birth length predicts stunting risk |

| OBJECTIVES | ENDPOINTS | JUSTIFICATION FOR ENDPOINTS |
|---|---|---|
| • Reduce LBW | Proportion of deliveries < 2.5kg | LBW is a risk factor for neonatal mortality |
| • Reduce neonatal mortality | Proportion of deaths < 28 days from delivery | Neonatal death rate is high in Sierra Leone. |
| • Reduce pre-term birth | Proportion of deliveries < 37 weeks | While not as severe as ePTB, still carries increased risk |
| • Reduce depression symptoms | Patient Health Questionnaire-9 score | As stated under primary objectives. |
| • Increase maternal and fetal DHA status | RBC-DHA in maternal and cord blood | Marker of accretion of DHA and adherence to intervention |
| • Improve sub-domains of cognitive development | MDAT gross motor, fine motor, language, social sub-domains | Assessment of individual domains to assess possible differential effects of intervention |
| • Increase post-term delivery | Proportion of deliveries > 42 weeks | Potential adverse effect of omega-3 supplementation in pregnancy |
| To determine whether adding 1g DHA/EPA + 550mg choline to a supplementary food provided to malnourished pregnant Sierra Leonean women **regardless of GA at enrollment** will:<br><br>• Prolong gestation<br>• Reduce early pre-term birth<br>• Improve birth weight and length<br>• Reduce LBW | | |
| Tertiary/Exploratory | | |
| To determine whether adding 1g DHA/EPA + 550mg choline to a supplementary food provided to malnourished pregnant Sierra Leonean women ≤ 30 weeks GA at enrollment will: | | |
| • Prolong gestation among subgroups by: | Gestational duration, days | Prior research has revealed possible differential effect of omega-3 by |

| OBJECTIVES | ENDPOINTS | JUSTIFICATION FOR ENDPOINTS |
|---|---|---|
| ○ Baseline DHA status<br>○ Baseline MUAC<br>○ Baseline gestational age<br>○ Parity<br>○ Maternal age | | maternal DHA status. Parity and maternal age are predictors of gestational duration. |
| • Reduce incidence of depression | PHQ-9 ≥ 9 | Cut-off commonly used to diagnose depression |
| • Increase placental weight | Placental weight (g) | Cause/marker of intrauterine growth restriction |
| • Increase birth chest and head circumference | Chest circumference (cm), head circumference (cm) | Birth chest circumference predicts neonatal and infant mortality |
| • Placental tissue changes | Placental weight | |
| • Increase gastrointestinal symptoms in mothers | Burping, constipation, diarrhea, vomiting | Patient-centered adverse effects |
| To determine whether provision of a novel cognitive behavioral therapy intervention among women with ante- or postnatal depression will: | | |
| • Prolong gestation | Gestational duration, days | |
| • Improve infant cognitive development | MDAT | |

### 4 STUDY DESIGN

#### 4.1 OVERALL DESIGN

We hypothesize that adding 500mg DHA, 500mg EPA, and 550mg choline to a supplementary food (M-RUSF+) provided to malnourished pregnant women in rural Sierra Leone will (1) prolong gestation and (2) improve offspring Malawi Developmental Assessment Tool global z-scores at 9 months of age compared with provision of a similar supplementary food but without DHA/EPA/choline (M-RUSF), and that (3) provision of a novel cognitive behavioral therapy (CBT) program to women with ante- or postnatal depression will reduce depressive symptoms.

This will be a 2x2 factorial, randomized, controlled, partially blinded superiority trial. All participants will be individually randomized, 1:1, to receive either M-RUSF+ or M-RUSF. Participants who are found to have PHQ-9 scores ≥ 9 will be individually randomized, 1:1, either to receive (intervention) or not receive (control) a novel CBT program. The table below summarizes the structure.

| Depression status | CBT vs. none | M-RUSF+ | M-RUSF |
|---|---|---|---|
| PHQ-9 ≥ 9 | CBT | | |
| | No CBT | | |
| PHQ-9 < 9 | No CBT | | |

The M-RUSF+ vs. M-RUSF comparison will be fully blinded. Care was taken in food design to minimize differences in taste, aftertaste, and texture between the foods. Outcomes assessors will be blinded. In the CBT vs. none comparison, participants and therapists will not be blinded. Outcomes assessors will be blinded, as PHQ-9 assessments will take place at clinic and not by therapy staff. In both cases, the data analysis team will be blinded.

M-RUSF and M-RUSF+ will be composed of whey protein isolate, skim milk powder, peanuts, palm oil, pearl millet, sugar, and multiple micronutrient mix. M-RUSF+ will also contain 1.7g of fish oil (500mg DHA+ 500mg EPA) and 550mg choline given as choline chloride. CBT will involve 6 bi-weekly 45-minute sessions.

This trial will take place across 30-40 government-run antenatal clinics in Pujehun and Bo Districts, Southern Sierra Leone. Total clinic number will depend on rate of accrual.

#### 4.2 SCIENTIFIC RATIONALE FOR STUDY DESIGN

M-RUSF functions as a placebo control. It contains the same ingredients as M-RUSF+ except without DHA/EPA/choline. Care will be taken in food design to minimize differences in taste, after-taste, and texture. Because these ingredients cost money, the trial is designed to demonstrate superiority. CBT will not have a control. Because CBT takes time and effort, the comparison is designed to demonstrate superiority.

#### 4.3 JUSTIFICATION FOR DOSE

The Institute of Medicine does not set a daily recommended intake for DHA in pregnancy. 200mg / day is recommended by some expert groups (Simopoulos 2000, FAO WHO 2004, Koletzko 2007).[23,24] Trials that have found an effect of omega-3 PUFA supplementation on gestational duration / early preterm birth have provided >600 DHA. These include Olsen 1992, DOMInO, KUDOS, and ADORE.[20,25-27] There is uncertainty as to the amount of omega-3 PUFA consumed across Sierra Leone, with likely substantial variability primarily driven by differential access to fish. We expect malnourished women to have low intake and levels of omega-3 PUFA. We will provide 500mg/500mg DHA/EPA, expecting that significant amounts of EPA will be converted to DHA but still allowing the opportunity for EPA supplementation to improve depressive symptoms (secondary outcome).

There is uncertainty as to the ideal recommended dose of choline for pregnant women. The uncertainty is greater still in the setting of undernutrition. The Institute of Medicine recommends pregnant women to consume 450mg choline daily, on average. Subsequent to this recommendation, research suggests that the 25mg increase in recommended dose for women who are pregnant (from 425mg for non-pregnant women) is insufficient to account for greater physiologic demands for choline's functions during pregnancy.[28] A 2022 trial found that 550mg of choline improved hepatic DHA export and biomarkers of DHA status among pregnant patients consuming DHA.[16] We expect malnourished women in Sierra Leone to have low intakes and levels of choline, as the main sources are animal-source foods including milk and eggs. Thus, recognizing the lack of certainty around what constitutes an adequate dose, we will supplement 550mg of choline daily, which is much lower than doses with documented toxicity.

Albendazole (400mg), azithromycin (1g x2), and sulfadoxine/pyrimethamine (1,500mg/75mg monthly) will be provided at standard doses for use in pregnancy. WHO recommends 400mg albendazole after the first trimester in endemic regions, which includes Sierra Leone. WHO recommends at-least monthly SP at 1,500mg/75mg dose starting in the second trimester in endemic regions, which includes Sierra Leone. These doses were used in our prior RCT among malnourished pregnant women in Sierra Leone without issue.

Azithromycin is not guideline-recommended during pregnancy absent a specific indication. It has, however, been given in the second and third trimesters of pregnancy in multiple RCTs, 7 of which took place in low- and middle-income countries and included >1,200 participants each.[6] Several identified improvements in birth weight, LBW, and neonatal mortality, including MamaDutasi. Given the success of this study, and in light of prior positive findings, we will provide 1g of azithromycin in the second and third trimesters to all participants. The 1g dose is routine.

#### 4.4 END OF STUDY DEFINITION

A participant is considered to have completed the study if she has completed all phases of the study including the last visit as shown in the Schedule of Activities (SoA), Section 1.3.

The end of the study is defined as completion of the last visit or procedure shown in the SoA in the trial globally.

### 5 STUDY POPULATION

#### 5.1 INCLUSION CRITERIA

All women ≥ 13 years of age who present to a participating antenatal clinic, think they are pregnant, and express a willingness to undergo screening for the trial, will undergo screening. This will first include MUAC and BMI assessment. If a participant meets MUAC/BMI criteria for enrollment, they will undergo fundal height and ultrasound assessment. If they have a singleton pregnancy, they will be invited to undergo full inclusion/exclusion criteria assessment, consent, and randomization. For girls < 16 years of age, consent will require guardian/parent.

In order to be eligible to participate in the M-RUSF+ vs. M-RUSF element of the study, an individual must meet all of the following criteria:

1. Provision of signed (or thumb-printed) and dated informed consent form
   - 1a. Women ≥ 16 years of age will be allowed to consent for themselves
   - 1b. Women < 16 years of age must provide assent and a parent or guardian must provide consent
2. Stated willingness to comply with all study procedures and availability for the duration of the study, including no plan to move from the catchment area of a participating clinic
3. ≥ 13 years of age
4. Pregnant
5. Mid-upper arm circumference ≤ 23 cm or body-mass index < 18.5

In order to be eligible to participate in the CBT vs. no CBT element of the study (factorial design with the above), an individual must be enrolled in the M-RUSF+ vs. M-RUSF study and meet the following criteria:

1. Provision of signed (or thumb-printed) and dated informed consent form
   - 1a. Women ≥ 16 years of age will be allowed to consent for themselves
   - 1b. Women < 16 years of age must assent and a parent or guardian consent must provide consent
2. Stated willingness to comply with all study procedures and availability for the duration of the study, including no plan to move from the catchment area of a participating clinic
3. Patient Health Questionnaire-9 score ≥ 9

All malnourished pregnant women presenting to participating antenatal clinics, regardless of inclusion/exclusion criteria (aside from allergy to food/medications), will be offered the entire package of care included in the clinical trial.

#### 5.2 EXCLUSION CRITERIA

An individual who meets any of the following criteria will be excluded from participation in this study:

1. Participation in a concomitant supplementary feeding program
2. Known allergy to components of intervention or control study food or medications

3. Known gestational diabetes
4. Hypertension
5. Severe anemia, or other condition requiring immediate hospitalization

#### 5.3 LIFESTYLE CONSIDERATIONS

There will be no special lifestyle recommendations for this trial.

#### 5.4 SCREEN FAILURES

Screen failures are defined as participants who undergo screening but are not enrolled in the study. Most commonly, this is because they do not meet inclusion/exclusion criteria. Screen failure details will be collected throughout the trial to meet the Consolidated Standards of Reporting Trials (CONSORT) publishing requirements and to respond to queries from regulatory authorities.

Individuals who do not meet the criteria for participation in this trial (screen failure) because of MUAC/BMI above cut-offs for inclusion or age <13 years may be rescreened. Individuals who do not meet criteria for participation because of condition requiring immediate hospitalization may be rescreened.

#### 5.5 STRATEGIES FOR RECRUITMENT AND RETENTION

The target sample size is 1600. All participants will be malnourished, pregnant Sierra Leoneans, who would meet several criteria for vulnerability and under-represented status in clinical trials. In addition, we will include adolescent pregnant persons ≥ 13 years of age.

We anticipate accruing 70-80 participants / month. 30-40 government-run antenatal clinics in Bo and Pujehun Districts, Sierra Leone, will be included as sites of screening, enrollment, and trial operations. These clinics will be chosen in agreement with the District Medical Officers for each district and based on recruitment rates seen in the prior RCT by our group (Pujehun District) or census of pregnant women over the year preceding trial initiation (Bo District). In anticipation of the trial, clinic-catchment and village-level sensitization will be done by trial staff via meetings in local chiefs and elders, community health workers, and outreach visits to villages.

Participants may be involved in the trial for up to 16 months. During pregnancy, retention will be encouraged with bi-weekly visits at antenatal clinic, at which time health assessments will be done and supplementary food, recommended prenatal care, and study medications will be disbursed. Our prior experience with a similar study in a similar population suggests that receipt of food and medications will be a strong means of retention. We will also obtain directions from clinic to home for each participant to allow for visits by community health workers in the event of missed clinic visits (see **section 7.3**). We will obtain telephone numbers for the participant and a neighbor/family member. We will provide pre-paid telephone units with instructions to call clinic staff near the time of delivery. These measures are designed to increase retention and meet trial procedures for post-delivery assessments. In addition, participants will be given gifts of appreciation at enrollment, delivery, and all post-partum visits. Gifts are generally cloth wraps used for many purposes by women in Sierra Leone. Gifts will be given at each post-partum visit because participants will no longer be receiving food or medication after delivery, and thus incentives are required to maintain retention leading up to evaluation of the co-primary outcome MDAT at offspring 9 months of age.

### 6 STUDY INTERVENTION

#### 6.1 STUDY INTERVENTION(S) ADMINISTRATION

##### 6.1.1 STUDY INTERVENTION DESCRIPTION

Undernourished pregnant women are recommended by the WHO to receive a balanced energy and protein supplement. All pregnant women are recommended by WHO to receive daily iron (30-60mg of elemental iron) and folic acid supplementation (0.4mg). In populations with low dietary calcium intake, 1.5-2g per day of oral elemental calcium is recommended. There is strong evidence for multiple micronutrient supplementation in pregnancy. Otherwise, there are no international guidelines for nutritional supplementation in undernourished pregnant women.

We have modeled the intervention and control supplementary foods on those used in two previous RCTs by our group treating undernourished pregnant women in Malawi and Sierra Leone. Both foods were well-accepted by participants. A formal acceptability trial will be undertaken prior to main trial initiation to assess the acceptability of both M-RUSF+ and M-RUSF.

Both foods will be balanced protein-energy supplements and contain recommended micronutrients. Both were designed at the Food Laboratory of Mark Manary (Trial PI) at Washington University in St. Louis. Ingredients are listed in the table below. The micronutrient premix included in the RUSFs will provide the same quantities of micronutrients as the UNICEF/WHO/United Nations multiple micronutrient supplement for pregnant and lactating women (UNIMMAP), with additional calcium and magnesium.

| **Ingredient, g/100g** | **M-RUSF+** | **M-RUSF** |
|---|---|---|
| Pearl millet | 7.5 | 7.5 |
| Non-fat dry milk | 21.5 | 21.5 |
| Whey protein isolate | 6.8 | 6.8 |
| Palm oil | 23.8 | 26.3 |
| Brown sugar | 18 | 18 |
| Peanut | 15.9 | 15.9 |
| Fish oil (Polaris) | 1.7 | 0 |
| Choline chloride | 0.825 | 0 |
| Multiple micronutrient premix (DSM) | 4 | 4 |

The nutrient content of both foods is listed in the table below.

| **Nutrient** | **M-RUSF+, 100g** | **M-RUSF, 100g** | **Recommended daily allowance[29-31]** |
|---|---|---|---|
| Energy, Kcal | 530 | 540 | |
| Protein, g | 18.7 | 18.7 | |
| Fat, g | 33.7 | 34.5 | |
| n6 Polyunsaturated fatty acids, g | 6.5 | 6.5 | |
| n3 Polyunsaturated fatty acids, g | 1.3 | 0.3 | |
| Vitamin A, mg | 770 | 770 | 770 |

| | | | |
|---|---|---|---|
| Vitamin B1/Thiamine, mg | 2.8 | 2.8 | 1.4 |
| Vitamin B2/Riboflavin, mg | 2.8 | 2.8 | 1.4 |
| Vitamin B3/Niacin, mg | 32 | 32 | 18 |
| Vitamin B6, mg | 3.8 | 3.8 | 1.9 |
| Vitamin B12, mg | 5.2 | 5.2 | 2.6 |
| Folic acid, mg | 500 | 500 | 400 |
| Vitamin C, mg | 170 | 170 | 85 |
| Vitamin D, mg | 30 | 30 | 15 |
| Vitamin E, mg | 30 | 30 | 15 |
| Iron, mg | 30 | 30 | 27 |
| Zinc, mg | 22 | 22 | 11 |
| Calcium, mg | 1600 | 1600 | 1000 |
| Chromium, mg | 60 | 60 | 30 |
| Copper, mg | 2000 | 2000 | 1000 |
| Iodine, mg | 300 | 300 | 220 |
| Magnesium, mg | 300 | 300 | 350 |
| Phosphorus, mg | 687.5 | 687.5 | 700 |
| Potassium, mg | 892.7 | 892.7 | 4700 |
| Selenium, mg | 120 | 120 | 60 |

Both foods are light-brown-colored peanut-based pastes. They will be produced at the Project Peanut Butter (PPB) factory in Freetown, Sierra Leone. They will be nitrogen vacuum-sealed in foil sachets. The sachets will be identical in appearance aside from a sticker with a color signifying study group (M-RUSF+ vs. M-RUSF), the code for which will only be known by the staff responsible for packaging.

Each batch of M-RUSF+ and M-RUSF will meet UNICEF specifications for aflatoxin, Enterobacter sp., and Salmonella contamination. Boxes of sachets will be kept in a PPB storage room that is maintained at < 25°C until transfer to the research team for use at clinic.

The novel CBT program will consist of 6 hour-long sessions. Below is a summary of what might occur during a treatment program along with a few examples, though each treatment plan is tailored to the individual at hand by the counselor and participant. Beginning in session 1, a key goal is to create an enabling and empowering environment for women to talk openly and with assurance of confidentiality. Session 1 is also devoted to problem identification. This often involves a degree of unburdening, where the counselor's primary job is to listen, observe, and encourage sharing. In addition, the counselor will help the client to probe triggers for depressive symptoms as well as thought patterns and behaviors associated with psychological distress. Through so doing, they will attempt to identify targets for therapy and action. Sessions 2-3 often focus on generating potential solutions and choosing one or several for the participant to try. Between sessions, participants will be given so-called "homework," often involving small, manageable assignments. For instance, if a prominent issue is that the participant is feeling lonely or isolated, the counselor might encourage them to think through possible family, friend, or religious sources of support; for instance, to identify 5 such possibilities, reach out to those individuals to make a time to meet, and meet with them. Sessions 4 and 5 tend to focus on evaluation of attempted solutions and their impacts on maladaptive thoughts, behaviors, and signs/symptoms of depression. Homework assignments may be modified. Session 6 is often devoted to integration in two ways. First, our pilot experience has shown that isolation/shame/relationship challenges often factor heavily into ante- and post-natal depression in Pujehun District. For adolescents, this could mean they have dropped out of school, for instance, or been shunned by their families for becoming pregnant. The counselors will work with participants on re-integrating into their community to prevent the isolation that may have contributed to their depression. Second, counselors and participants will work on integrating the learned solutions to avoid/terminate unproductive thoughts and behaviors into the participant's repertoire of tools to continue treatment and prevent recurrence. The pilot program also identified group sessions as helpful for individuals who wish to engage in them. Therefore, all participants randomized to CBT will be offered the chance to engage in group sessions rather than exclusively in 1-on-1 sessions. The control group will receive no treatment.

###### 6.1.2 DOSING AND ADMINISTRATION

The amounts of DHA, EPA, and choline (which compose the difference between intervention and control foods) are described in **section 6.1**. Dose justification is provided in **section 4.3**. Participants will begin receiving the supplementary foods on the day of enrollment/randomization and continue until the date of delivery. Participants will receive sufficient supply for 1 packet per day (100g, 5040 Kcal). Participants will be given a 2-week supply at each bi-weekly prenatal visit. Participants will be encouraged to consume their supplementary food every day. Participants will be instructed that the food requires no preparation or modification prior to consumption directly from the sachet, though they may mix it into a porridge, for instance. If a participant misses a study visit, they will not be able to receive their supplementary food for the following 2 weeks.

The earliest anticipated enrollments will occur at 10 weeks' gestation. Thus, we estimate the longest duration of supplementary feeding to be 30-32 weeks. We estimate the shortest duration of supplementary feeding to be 2 weeks, as GA is not a criteria for trial enrollment and early preterm birth is not uncommon in this population.

All participants will receive 400mg of albendazole on or after 13 weeks' gestation, 1g of azithromycin during the second and third trimesters (or only third, if enrolled during the third trimester), and 1,500 mg/75 mg of sulfadoxine/pyrimethamine (SP) every 4 weeks beginning at 13 weeks' gestation. Dose justifications are provided in Section 4.3. Each of these medicines will be given under direct observation at antenatal clinic.

Most CBT sessions will take place individually between study therapists and participants. In some cases, participants may wish to engage in group CBT sessions and, if possible, therapists will then work to set up group sessions for those interested.

##### 6.2 PREPARATION/HANDLING/STORAGE/ACCOUNTABILITY

###### 6.2.1 ACQUISITION AND ACCOUNTABILITY

M-RUSF+ and M-RUSF will be produced at the Project Peanut Butter (PPB) factory in Freetown, Sierra Leone. PPB is an NGO that has been operational in Sierra Leone since 2007. Mark Manary, the trial PI, is the founder and CEO of PPB. The study foods will undergo microbial and toxin testing as per UNICEF specifications. Boxes of study food will be delivered to the research teams in Pujehun and Bo Districts as needed. Chain of custody will be documented with signatures. Study foods will be disbursed to participants at antenatal clinic by dedicated research staff, who will document receipt including colored sticker signifying study group. Participants will be instructed to return all empty sachets the following clinic visit as a loose measure of adherence and to prevent litter from accumulating in study villages.

Albendazole, azithromycin, and SP will be obtained from registered pharmacies in Freetown, Sierra Leone. Insecticide-treated bednets will also be purchased in Freetown. Safe birth kits will be distributed by the study which include sterile tools to cut/clamp the umbilical cord and clean sheets to dry and wrap the newborn.

##### 6.2.2 FORMULATION, APPEARANCE, PACKAGING, AND LABELING

M-RUSF+ and M-RUSF formulations are detailed in **section 6.1.1**. Both foods will appear as light brown peanut-based pastes. Most ingredients have been used to produce therapeutic and supplementary foods at the PPB factory for nearly 10 years. The new ingredients are DHA, EPA and choline.

DHA/EPA have been added to therapeutic foods at PPB factories in Sierra Leone and Malawi since 2022. Our group performed formal testing of final product DHA content under various conditions and published this work in 2023.[32] In accordance with these findings, the fish oil will be added toward the end of the production process to reduce degradation/oxidation. Samples of the M-RUSF+ will be tested for DHA content quarterly during the trial to assess levels in the final product.

Choline is added to a variety of food products used worldwide. In order to maximize incorporation into the paste, it will be added at the first mixing step.

M-RUSF+ and M-RUSF will be packaged in identical foil sachets, aside from the presence of a sticker with a color indicating study group. The foods will be produced by PPB in Freetown, Sierra Leone.

Albendazole, azithromycin, and SP will be purchased locally in Freetown, Sierra Leone at registered pharmacies. There will likely be several manufacturers.

##### 6.2.3 PRODUCT STORAGE AND STABILITY

M-RUSF+ and M-RUSF will be packaged in vacuum-sealed metallized polyester sachets with a nitrogen flush that are also used to package RUTF to meet UNICEF specifications. These packets will be placed into cardboard boxes which will be stored in a cool, dark, dry storage room at the PPB factory in Freetown and kept at < 25°C. Extrapolating from similar products (RUTF, RUSF) produced by PPB using identical packaging, shelf-life in foil packets will exceed 2 years. Once opened, the contents are recommended to be consumed within 24 hours. Study foods will be disbursed within 9 months of production.

DHA content testing will be done on both fresh and stored (9 months) products to assess stability.

Albendazole, azithromycin, and SP will be kept in original bottles in dry storage rooms at ambient temperature at the research team headquarters in Bo and Pujehun Districts. All medications will be given within 6 months of purchase and preceding indicated expiration dates.

###### 6.2.4 PREPARATION

M-RUSF+ and M-RUSF will be ready for consumption when disbursed, requiring no further preparation or manipulation by study staff or participants. Participants will be told they are allowed to mix the study food into a porridge if that is their preference.

##### 6.3 MEASURES TO MINIMIZE BIAS: RANDOMIZATION AND BLINDING

**Randomization**
Participants will be randomized individually in a 1:1 ratio to M-RUSF+ vs. M-RUSF; among these participants, those with PHQ-9 ≥ 9 will be individually randomized in a 1:1 ratio to CBT vs. no CBT. A research team member will prepare the method of randomization for M-RUSF+ vs. M-RUSF as follows. There will be 6 colored stickers, 3 of which will indicate each study group. The stickers will adhere to a piece of paper that will allow for easy removal and re-application. The colors will be black, white, blue, yellow, green, and red. A research team member will collect 4 of each colored sticker (24 total) and 24 small opaque envelopes. They will place one sticker into each envelope, seal the envelopes, and place all 24 small envelopes into a larger opaque manilla envelope. 75 such larger envelopes will be prepared prior to study initiation and will be used one at a time. At the time of randomization, a participant will blindly reach in the larger opaque manilla envelope and withdraw one smaller opaque envelope, open it, and thereby choose their study group. The sticker will then be placed on their study card at two locations.

The method of randomization for CBT vs. no CBT will be similar, with 6 separate symbols, 3 to each group. 10 larger envelopes will be prepared in advance of the trial. After a participant randomizes themselves by choosing a symbol, this symbol will be stuck to their CBT visit card. A team member unblinded to the allocation code for CBT vs. no CBT will then instruct the participant on whether they are to receive CBT and, if so, the schedule for their sessions.

**Blinding**
<u>M-RUSF + vs. M-RUSF</u>
M-RUSF+ and M-RUSF will be packaged in identical foil sachets aside from a colored sticker, which will indicate study group. The two foods are identical in appearance and similar in texture and taste.

A staff member in the US responsible for scheduling M-RUSF+ and M-RUSF production at the PPB factory and scheduling delivery of boxes of food to research staff will be unblinded as to the color-group code. Staff members at the PPB factory responsible for food production and labeling the sachets will also be unblinded. None of these staff members will take part in the study otherwise, including interaction with participants, clinic visits, or outcomes assessments. They will keep the code locked.

All research clinic staff/outcomes assessors will remain blinded to M-RUSF+ vs. M-RUSF allocation throughout the trial. Staff responsible for disbursing study foods will do so by matching the color to which a participant was randomized (which will be attached to study cards) to the color on the sachets and boxes containing sachets but will not know the underlying allocation code. Three colors will be used per group to minimize the chance of accidental unblinding, e.g., if M-RUSF+ leads to greater levels of burping or constipation, the staff member may be able to link such symptoms to a color but will have more trouble identifying group membership of other colors. Following delivery (first postpartum visit and onward), new study cards will be made for all participants which will not contain their randomization group. Thus, M-RUSF+ vs. M-RUSF group membership will not be displayed from delivery until MDAT testing (end of trial).

Neurocognitive assessors will be blinded to M-RUSF+ vs. M-RUSF, as the colors to which participants were randomized will not be present on visit cards.

Staff responsible for database management and data analysis will remain blinded to M-RUSF+ vs. M-RUSF allocation, as randomization colors will be used in the database and the code will remain locked until all analyses have been completed.

CBT vs. no CBT

Neither participants nor therapists will be blinded to CBT vs. no CBT allocation. A clinic staff member on each team will know the code linking randomization symbol to CBT vs. no CBT so they can notify the participant whether they have been randomized to receive CBT and to schedule their first therapy visit if so. This team member will be designated in advance and will not take place in PHQ-9 assessments during the trial.

For the purposes of tracking their therapy, counselors will perform PHQ-9 assessments and function assessments periodically. These data will be collected and used for safety analysis but will not be used to assess efficacy of CBT vs. no CBT, as the counselors are not blinded. Instead, a nurse or counselor trained in PHQ-9 assessment and blinded to allocation will perform a PHQ-9 assessment 8 weeks after randomization into the CBT vs. no CBT trial. During the antenatal period, all participants (regardless of enrollment in the CBT vs. no CBT trial) will undergo bi-weekly PHQ-9 assessment by a nurse or counselor who will not know whether the participant is in the CBT vs. no CBT trial or which group they are allocated to, if so. During the postnatal period, all participants will undergo PHQ-9 assessments at their scheduled visits by a nurse or counsellor who again will not be aware of enrollment in or group allocation for the CBT vs. no CBT trial.

Neurocognitive assessors will be blinded to CBT vs. no CBT, as the symbols to which participants were randomized will not be present on visit cards.

#### 6.4 STUDY INTERVENTION COMPLIANCE

This trial has been designed to approximate practices that might happen were its results to be applied in resource-constrained settings with high rates of undernutrition in pregnancy. That is, malnourished pregnant women will be provided with food and encouraged to consume it, but this consumption will not be directly monitored to confirm receipt. Adherence to M-RUSF+ vs. M-RUSF will be assessed in several ways, however. First, participants will be asked to bring empty sachets back to clinic fortnightly, and these will be identified by group color and be counted. Second, we will enlist community health workers stationed at each antenatal clinic to visit participants at their homes at random (schedule made using a random number generator) approximately q6 weeks to document the colored sticker on a participant's sachets and the number of full sachets remaining on date of visit, which will then be compared to expectations based on their last study visit. Third, blood spots will be collected from participants at baseline and between 32-36 weeks' gestation for RBC-PUFA status, which will provide a sense of overall adherence.

Adherence to albendazole, azithromycin, and SP will be known because each medication will be given under direct observation at antenatal clinic.

Adherence to CBT will be known by documentation of visits by therapists.

#### 6.5 CONCOMITANT THERAPY

For this protocol, a prescription medication is defined as a medication that can be prescribed only by a properly authorized/licensed clinician. Medications to be reported on participant data collection forms on a bi-weekly basis are concomitant prescription medications, over-the-counter medications, and supplements.

Receipt of albendazole, azithromycin, and SP will be documented on study cards.

##### 6.5.1 RESCUE MEDICINE

Not applicable.

### 7 STUDY INTERVENTION DISCONTINUATION AND PARTICIPANT DISCONTINUATION/WITHDRAWAL

#### 7.1 DISCONTINUATION OF STUDY INTERVENTION

Discontinuation from supplementary food receipt or CBT does not mean discontinuation from the study, and remaining study procedures should be completed as indicated by the study protocol. If a clinically significant finding is identified after enrollment, the first priority will be clinical management of the participant, including a triage decision. This decision will be made by the most clinically experienced staff member present, often a nurse-midwife. The research associate will notify the principal investigator within 3 days to determine if any change in study management is needed. Any new clinically relevant finding will be reported as an adverse event (AE). The two foreseeable reasons for intervention discontinuation are participant choice and allergy to M-RUSF+ / M-RUSF. The data to be collected at the time of study intervention discontinuation will include the following:

- Reason for discontinuation
- Maternal anthropometry
- Physical examination and symptom evaluation (e.g., rash, abdominal discomfort, trouble breathing)
- Need for hospitalization
- Outcome of hospitalization, if needed

Participants who discontinue the intervention will be encouraged to remain in the study for follow-up. If they agree, they will undergo the same data collection as those who remain on the intervention.

In the case of participants who have been diagnosed with depression (whether randomized to CBT or not), the same procedures will be undertaken as above. In the event of active suicidal or homicidal ideation, the participant will be offered transport to the Pujehun District Hospital and the counselor's clinical supervisor will be informed to advise and direct care. The research associate will notify the principal investigator within 3 days.

#### 7.2 PARTICIPANT DISCONTINUATION/WITHDRAWAL FROM THE STUDY

Participants are free to withdraw from participation in the study at any time upon request. An investigator may discontinue or withdraw a participant from the study for the following reasons:

- If any clinical adverse event (AE), laboratory abnormality, or other medical condition or situation occurs such that continued participation in the study would not be in the best interest of the participant
- If the participant meets an exclusion criterion (either newly developed or not previously recognized) that precludes further study participation
- Participant defaults, i.e. misses 3 consecutive study visits

The reason for participant discontinuation or withdrawal from the study will be recorded on the participant data card. Individuals who sign the informed consent form, are randomized and receive the study intervention, and subsequently withdraw, or are withdrawn or discontinued from the study, will not be replaced.

#### 7.3 LOST TO FOLLOW-UP

A participant will be considered lost to follow-up (defaulted) if they fail to return for 3 consecutive study visits and are unable to be contacted by the study site staff.

The following actions will be taken if a participant fails to return to the clinic for a required study visit:

- Staff will attempt to contact the participant via telephone, send a message with their next study visit date via a community health worker (CHW), ask whether the woman remains pregnant, reschedule the missed visit, counsel the participant on the importance of maintaining the assigned visit schedule, and ascertain if the participant wishes to and/or should continue in the study. CHWs will receive a small gift for any successful participant tracing or information obtained via their visit.
- Before a participant is deemed lost to follow-up, the investigator or designee will make efforts to regain contact with the participant: at least 3 telephone calls, asking a CHW to visit the participant at their home 2 times, and taking a trip with the research team to the participant's documented address after 3rd consecutive missed visit. These contact attempts will be documented in the participant's study file.
- Should the participant continue to be unreachable, he or she will be considered to have withdrawn from the study with a primary reason of lost to follow-up.

### 8 STUDY ASSESSMENTS AND PROCEDURES

#### 8.1 EFFICACY ASSESSMENTS

**Screening: M-RUSF+ vs. M-RUSF**

There will be two settings for screening. The first, which will afford partial screening/sensitization, will be done by antenatal clinic nurses and community health workers (CHWs). The study team will disburse mid-upper arm circumference (MUAC) tapes at each participating antenatal clinic for use at clinic. Nurses and CHWs at each clinic will be informed to offer screening with MUAC measurement to any woman who thinks she may be pregnant. All women who think they are pregnant, regardless of MUAC, will be recommended to attend the next antenatal clinic day. If a woman is found to have MUAC ≤ 23.0 cm, they will be informed of the trial and possibility of their enrollment pending further evaluation. If possible, antenatal clinic staff may also pursue BMI measurements, in which case BMI < 18.5 would trigger the same discussion. All women who think they may be pregnant will be encouraged to visit antenatal clinic. Those with low MUAC will be informed of the trial and possibility for enrollment if all criteria are met.

Screening for trial enrollment will take place at bi-weekly antenatal clinic visits by research staff, primarily Sierra Leonean nurses, a full-time research associate, and US-trained MDs when they are present in Sierra Leone. Women who think they may be pregnant will be asked whether they would like to be screened for possible trial enrollment, a process which will involve anthropometry, brief history questions, fundal height measurement, and ultrasonography for estimated gestational age. Women who consent will first be asked their age. If they are < 13 years of age, they will not be eligible for the trial. If they are < 16 years of age, they must have a parent/guardian willing to consent for them. Next, MUAC and BMI measurements will be done by research staff. If potential participants meet eligibility criteria by MUAC or BMI, they will undergo fundal height measurement. This measurement will be used to guide the transabdominal ultrasound procedure, specifically probe measurement depth.

An ultrasound interpretation program imbedded within the Butterfly software will then be used to assess for singleton vs. other numbered pregnancies. All ultrasonography will be done by research staff who have received training in detection of singleton pregnancies and use of gestational age estimation software (as detailed elsewhere). Women with singleton pregnancies will be invited to continue screening.

Following successful GA estimation of a singleton pregnancy, exclusion criteria will be assessed: involvement in a separate supplementary feeding program, allergy to intervention components, history of gestational diabetes, blood pressure measurement, and general clinical assessment for pregnancy complications requiring immediate hospitalization, such as signs and symptoms of pre-eclampsia or severe anemia. If their blood pressure is > 140/90, they will not be eligible for enrollment and will be referred for further care. If they have a clinical indication for immediate hospitalization, they will not be eligible for enrollment at that visit and will be offered transport to Pujehun District Hospital accompanied by our research nurses and field staff.

If they meet all study inclusion/exclusion criteria, full discussion of trial procedures and formal consent process will be done. If the woman consents (or, if <16 years of age, assents with caregiver/guardian consent), they will then undergo randomization, as described elsewhere. Women who meet enrollment criteria and consent to participation will be enrolled, with interventions initiated, that same day. This includes study food and medications with timing detailed elsewhere in this protocol.

If the woman does not meet criteria by MUAC or BMI, they will be encouraged to continue attending antenatal clinic and may be screened on subsequent clinic days; if they meet criteria in the future, the above procedures will be done. Similarly, if a woman is < 13 years of age, she will be encouraged to continue attending antenatal clinic and may be screened on subsequent clinic days. Women who are referred for hospitalization may also return to antenatal clinic after recovery for repeat screening.

**Screening: CBT vs. no CBT**
All participants in the M-RUSF+ vs. M-RUSF comparison will undergo PHQ-9 testing bi-weekly throughout their pregnancy and at each postpartum study visit. If their PHQ-9 ≥ 9 at any time, they will be offered enrollment into the CBT vs. no CBT comparison. If they meet all inclusion/exclusion criteria and consent (or assent + parental/guardian consent if <16 yo) to participation, they will then be randomized. CBT will start within the week following randomization.

**Efficacy assessments**
**Maternal**

- Gestational duration: this will be determined by adding the estimated gestational age (GA, days) at enrollment to the time (days) between enrollment and date of delivery. Enrollment GA estimation ultrasonography will be done by trained research staff nurses using Butterfly probes and the Butterfly Gestational Age Tool, which uses an algorithm (not commercially available as of this writing) imbedded within the Butterfly application. This program was developed by The Fetal Age Machine Learning Initiative (FAMLI), an ongoing project chaired by Jeffrey Stringer, MD, of the University of North Carolina. The algorithm was validated in a large Zambian cohort, including when used by untrained sonographers performing blind transabdominal sweeps.[33] Our Sierra Leonean research nurses will be formally trained prior to trial initiation by FAMLI staff from Zambia who worked on development of this tool. The GA estimation ultrasound protocol is detailed in an SOP elsewhere. Briefly, first, fundal height will be measured and entered into the program to determine the number of lateral sweeps necessary. The Tool will then guide the user to conduct cranio-caudal and lateral sweeps. Each sweep is expected to last 10 seconds; if the sweep is done too quickly or slowly, the program automatically prompts the user to re-do the sweep. For each sweep, the app directs the user on location and direction, and counts down from 10 seconds. First, 5 cranio-caudal sweeps will be done, starting at the midline at the pubis and ending at the xyphoid. Next, 2 cranio-caudal sweeps are done to the left of midline, and then 2 to the right, with each sweep overlapping ~50% which its nearest neighbor. The Tool will then guide the user to perform lateral sweeps, beginning just above the pubic symphysis, collecting from the left lateral edge of the uterus to the right lateral edge of the uterus and repeating sweeps superiorly with 1 probe's distance between. The user may be guided to conduct 3-4 sweeps depending on fundal height. After the last sweep is complete, the user selects "Calculate" on the Tool. If the sweeps were sufficient, the Tool will calculate an estimated GA, which the user then saves and transcribes to the participant's data card. Once staff can access WiFi, the studies (video loops of each sweep) and results will be uploaded to a secure cloud-based database. Every ultrasound video collected will be uploaded to a secure storage cloud within a week of collection.
- Malawi Developmental Assessment Tool: testing will take place according to the SOP detailed elsewhere. Briefly, neurocognitive assessors will undergo training prior to trial initiation. At the 9mo postpartum visit, these neurocognitive assessors will assess gross motor, fine motor, and

language development via direct testing of the infant, and social development via caregiver interview, across 136 items total. Each item is scored as pass, fail, or NA. NA is used when a child does not engage sufficiently to allow a determination. As per testing recommendations, six consecutive fail determinations will trigger an automatic jump to the next domain of testing. This testing will be done by trained neurocognitive assessors in the participant's native language. Data will be collected on tablets with secure storage cloud upload. A program developed for the MDAT automatically calculates age-adjusted z-scores globally and for each sub-domain.

- PHQ-9: Standard 9-item questionnaire will be used wherein respondents are asked to estimate the number of days in the preceding 2 weeks that they have been bothered by each item. PHQ-9 testing will be done by research staff nurses in participant native language and with pictorial guides that have been developed and successfully piloted for use in Pujehun District, Sierra Leone, by the trial PI. Scores will be recorded on participant data sheets. In addition, PHQ-9 assessments will be done periodically by CBT counselors as clinically indicated, though these scores will not be used for outcomes assessment, but rather the guide CBT.
- Maternal MUAC: MUAC will be measured in duplicate by 2 study staff to the nearest 1mm with a standard insertion measuring tape (TALC, St. Albans, UK), according to standard procedures.
- Maternal weight: weight will be measured in duplicate with a Seca 803 digital scale to the nearest 100g.
- Maternal height: height will be measured in duplicate with a Seca 213 stadiometer to the nearest 1mm.
- Fundal height: fundal height will be measured in duplicate to the nearest 5mm using a non-elastic measuring tape.
- Blood spot: a blood spot will be taken at enrollment and at gestational age 32-36 weeks in selected subjects according to SOP detailed elsewhere.
- Cord blood: a cord blood sample will be collected from participants at selected centers as possible, according to SOP detailed elsewhere.
- Placental weight: the placenta will be weighed from participants at selected centers as possible, according to SOP detailed elsewhere.
- Saliva: saliva sample will be collected from selected participants for DHA assessment
- Offspring vital status. If found to be deceased, a standardized Verbal Autopsy Tool will be used.
- Infant MUAC: measured in duplicate by 2 study staff to the nearest 1mm with a standard insertion tape
- Infant weight: measured in duplicate using a Seca 334 digital scale
- Infant length: measured in triplicate to the nearest 2mm using a rigid length board
- Infant chest circumference: measured in duplicate to the nearest 1mm with a standard insertion tape
- Infant head circumference: measured in duplicate to the nearest 1mm with a standard insertion tape
- Infant mid-thigh circumference: measured in duplicate to the nearest 1mm with a standard insertion tape.

All maternal and infant anthropometry will be performed by trained research nurses, the full-time research associate, or US-trained MDs. Birth assessments will be done by a team designated to respond to notification of birth within 48 hours.

Participants will be notified of their anthropometric measurements, blood pressure, estimated gestational age, and fundal height measurements.

**Administration of trial interventions**
Participants will be provided with 2 weeks of their randomized study food prior to leaving clinic on their day of enrollment. They will be instructed on opening and administration of the study food, with particular attention given to avoiding sharing of the food amongst others. They will be counseled that this food has been designed to help them and their baby. On appropriate clinic days, they will be given albendazole, azithromycin, and/or SP under direct observation by research staff nurses. Timing of these is described elsewhere.

In addition to trial interventions, all participants will receive standard antenatal care as per Sierra Leone national guidelines. This care will be provided by Sierra Leone district medical staff. They will also receive safe birth kits.

#### 8.2 SAFETY AND OTHER ASSESSMENTS

**Safety Assessments**
- Participants will be asked about bleeding at each bi-weekly prenatal visit
- Gastrointestinal distress: participants will be asking about vomiting and diarrhea at each bi-weekly prenatal visit
- Participants will be asked about fever, rash, cough at each bi-weekly prenatal visit
- Maternal death
- Maternal prenatal hospitalization
- Post-term delivery (≥ 42 weeks gestation)
- Peripartum complication requiring transfer to facility with higher level of care
- Miscarriage
- Stillbirth
- Infant death

**Additional Assessments**
- Demography: maternal date of birth, age, level of education, # of children in household, # adults in household, father of coming child live with mother, # people sleep in same room as mother last night, household roof material, # radios/bicycles in home, house with electricity, cellphone in home, household water source, # times per day collecting water, use of pit latrine, animals sleep in home last night
- Health history: illness in last 2 months (none, diarrhea, pneumonia, TB, malaria), reduction in food intake during past 6 months, mother on TB treatment, other person in house on TB treatment, sleep beneath bed net, chew/sniff tobacco, kola nut use, alcohol consumption, pregnany history (#, # live births, age of youngest child, # miscarriages, # stillbirths), currently breastfeeding, currently taking supplements
- Household food insecurity access questionnaire
- Blood pressure: BP will be measured using a digital BP cuff. If elevated, BP will be re-checked using a manual sphygmomanometer for confirmation/refutation.
- Health data: last menstrual period (as estimated by mother), HIV test/status

#### 8.3 ADVERSE EVENTS AND SERIOUS ADVERSE EVENTS

##### 8.3.1 DEFINITION OF ADVERSE EVENTS (AE)

As adopted from 21 CFR 312.32 (a):

An adverse event means any untoward medical occurrence associated with the use of an intervention in humans, whether or not considered intervention-related.

##### 8.3.2 DEFINITION OF SERIOUS ADVERSE EVENTS (SAE)

As adopted from 21 CFR 312.32 (a):

An adverse event or suspected adverse reaction is considered "serious" if, in the view of the investigator, it results in any of the following outcomes: death, a life-threatening adverse event, inpatient hospitalization or prolongation of existing hospitalization, a persistent or significant incapacity or substantial disruption of the ability to conduct normal life functions, or a congenital anomaly/birth defect. Important medical events that may not result in death, be life-threatening, or require hospitalization may be considered serious when, based upon appropriate medical judgment, they may jeopardize the participant and may require medical or surgical intervention to prevent one of the outcomes listed in this definition.

##### 8.3.3 CLASSIFICATION OF AN ADVERSE EVENT

###### 8.3.3.1 SEVERITY OF EVENT

For adverse events, the following guidelines will be used to describe severity.

- **Mild** – Events require minimal or no treatment and do not interfere with the participant’s daily activities.
- **Moderate** – Events result in a low level of inconvenience or concern with the therapeutic measures. Moderate events may cause some interference with functioning.
- **Severe** – Events interrupt a participant’s usual daily activity and may require systemic drug therapy or other treatment. Severe events are usually potentially life-threatening or incapacitating.  Of note, the term “severe” does not necessarily equate to “serious”.

###### 8.3.3.2 RELATIONSHIP TO STUDY INTERVENTION

On the basis of prior trial data as well widespread use of prenatal fish oil supplementation and presence of comparable amounts of choline in widely consumed foods, there are no anticipated serious adverse events associated with addition of fish oil/choline to the supplementary food. Similarly, while CBT can lead to short-term distress during therapy, serious adverse events are also not anticipated for this technique that is widely used for management of ante- and post-natal depression. In a pilot program ongoing in Sierra Leone that has included >500 women with ante- or post-natal depression, there have been no cases of SI or HI that developed during CBT. In addition, and in line with prior clinical trials of prenatal fish oil supplementation, while non-serious adverse events typically associated with pregnancy will be monitored, recorded, and reported at the bi-monthly data safety monitoring meetings, they will not be actively assessed for relation to study intervention.[25]

As detailed below in several sections, on a monthly basis, the study monitor will compile all adverse events by study group. If an imbalance is identified, they will inform the principal investigator within 3 days. The principal investigator will then review the unblinded aggregate data to make a determination about potential relatedness to the intervention using the categories below. They will then notify SLESRC and the Wash U IRB within 24 hours. The principal investigator will then make a decision as to whether the trial can proceed.

- **Definitely Related** – There is clear evidence to suggest a causal relationship, and other possible contributing factors can be ruled out. The clinical event occurs in a plausible time relationship to study intervention administration and cannot be explained by concurrent disease or other drugs or chemicals. The response to withdrawal of the study intervention should be clinically plausible. The event must be pharmacologically or phenomenologically definitive, with use of a satisfactory rechallenge procedure if necessary.
- **Probably Related** – There is evidence to suggest a causal relationship, and the influence of other factors is unlikely. The clinical event occurs within a reasonable time after administration of the study intervention, is unlikely to be attributed to concurrent disease or other drugs or chemicals, and follows a clinically reasonable response on withdrawal. Rechallenge information is not required to fulfill this definition.
- **Potentially Related** – There is some evidence to suggest a causal relationship. However, other factors may have contributed to the event (e.g., the participant's clinical condition, other concomitant events). Although an AE may rate only as "possibly related" soon after discovery, it can be flagged as requiring more information and later be upgraded to "probably related" or "definitely related", as appropriate.
- **Unlikely to be related** – A clinical event whose temporal relationship to study intervention administration makes a causal relationship improbable (e.g., the event did not occur within a reasonable time after administration of the study intervention) and in which other drugs or chemicals or underlying disease provides plausible explanations (e.g., the participant's clinical condition, other concomitant treatments).
- **Not Related** – The AE is completely independent of study intervention administration, and/or evidence exists that the event is definitely related to another etiology. There must be an alternative, definitive etiology documented by the clinician.

###### 8.3.3.3 EXPECTEDNESS

There will be two mechanisms of expectedness assessment, including both trial PI Mark Manary and the Independent Monitoring Safety Committee (IDMC). AEs and SAEs will be considered unexpected if the nature, severity, or frequency of the event is not consistent with the risk information previously described for the study intervention.

##### 8.3.4 TIME PERIOD AND FREQUENCY FOR EVENT ASSESSMENT AND FOLLOW-UP

The occurrence of an AE or SAE may come to the attention of study personnel during study visits (bi-weekly prenatal and 5 postnatal visits), either solicited or unsolicited, or via report from antenatal clinic staff regarding events that occurred between study visits.

All AEs including local and systemic reactions not meeting the criteria for SAEs will be captured on the participant's data card. All AEs not typically associated with pregnancy in the clinician's judgement will undergo further assessment. This can include symptoms, signs, severity, duration, or other aspects of the AE not typically associated with pregnancy. Information to be collected for these AEs includes event description, duration, clinician's assessment of severity, possible relation to study product (food, medication), and time of resolution/stabilization of the event. All AEs occurring while on study must be documented appropriately regardless of relationship. All AEs will be followed to adequate resolution.

Any medical condition that is present at the time that the participant is screened will be considered as baseline and not reported as an AE. However, if the study participant's condition deteriorates at any time during the study, it will be recorded as an AE.

Changes in the severity of an AE will be documented to allow an assessment of the duration of the event at each level of severity to be performed. AEs characterized as intermittent require documentation of onset and duration of each episode.

All reportable events will be recorded with start dates (may be approximate) occurring any time after informed consent is obtained until the last day of study participation.

In addition to monitoring in the normal course of clinical trial operations, the IDMC will be responsible for reviewing AE and SAE data yearly.

##### 8.3.5 ADVERSE EVENT REPORTING

AEs typically associated with pregnancy that are occurring in an expected manner in the clinician's judgment will not be actively reported. If an AE typically associated with pregnancy differs in any way deemed significant by the clinician, such will be documented on the study form and discussed with at least a nurse-midwife and the research associate to determine if clinical management steps need to be taken. If the research team has concerns regarding the safety of continued intervention receipt, they will stop the intervention(s) and must then notify the study monitor and PI of the issue and their actions within 24 hours. The study monitor will analyze data collected up to that point to determine if the AE might be occurring at greater-than-expected rates or at greater rates in one or several groups compared to others. They will report to the PI, who will then decide how to proceed in consultation with nurse-midwife team.

If any research team member suspects AEs to be occurring at rates higher than anticipated or in a/several study groups more than others, they will notify the research associate, who will notify the study monitor within 3 days. The study monitor will investigate and report to the PI, who will decide how to proceed. Otherwise, on a monthly basis, the study monitor will compile AEs by study group. If rates are unbalanced or higher than anticipated based on comparison to prior trial data, they will notify the PI within 3 days and the PI will evaluate the unblinded aggregate summary data. The PI will notify SLESRC, PBSL, and WUSTL IRB and make a determination about how to proceed.

The IDMC will receive information on all reportable adverse events (events that are unexpected and related or probably related to the interventions) within 5 working days. The IDMC will independently review AEs at least yearly. The IDMC will generate a report on AEs for the PI, SLESRC, PBSL, and WUSTL IRB. Through its reviews of the study, the IDMC will determine whether cumulative data indicates the need to change the research design, to modify information presented to participants, or to terminate the project. Any action taken to suspend or terminate the project will be reported to the PI, SLESRC, PBSL, and WUSTL IRB. The IDMC will evaluate the final study manuscripts to assure results are fairly presented and conclusions are appropriate.

##### 8.3.6 SERIOUS ADVERSE EVENT REPORTING

In the event of an SAE, research staff must notify the research associate within 24 hours. The RA will attempt to gather information regarding the SAE. They will complete the SAE report form as outlined by the Pharmacy Board of Sierra Leone. They will send this report to the study monitor and PI within 24 hours, and to the PBSL within 48 hours. The study monitor will assess rates of this SAE in total and between groups and, if imbalance or concern for greater-than-expected rates are found, they will notify the PI who will notify SLESRC, PBSL, and WUSTL IRB and make a determination about how to proceed.

If any research team member suspects SAEs to be occurring at rates higher than anticipated or in participants within a study group more than others, they will be expected to notify the research associate, who will be required to notify the study monitor and principal investigator within 24 hours and PBSL within 48 hours. The study monitor will investigate by compiling SAE rates overall and by group and report to the PI within 24 hours, who will decide how to proceed. SAE rates irrespective of group will be compared to those identified in the 2019-2021 maternal undernutrition trial that took place amongst a similar population and with similar interventions in an effort to detect higher than anticipated SAE rates.

On a monthly basis, the study monitor will compile SAEs. If rates are higher than expected (compared to 2021 study), the study monitor will notify the principal investigator within 24 hours. The PI will notify SLESRC, PBSL, and the WUSTL IRB and make a determination about how to proceed. In addition, the study monitor will compare rates of SAEs between study groups. If rates are unbalanced, they will notify the PI within 24 hours and the PI will evaluate the unblinded summary data. The PI will notify SLESRC and WUSTL IRB and make a determination about how to proceed.

Follow-up reports on SAEs will be made and presented to SLESRC, PBSL, and WUSTL IRB when there is a change in the severity of SAE, when there is a new development on a previously reported SAE, or when the SAE resolved.

In the case of fatalities, if a formal autopsy is completed, this report will be saved and presented to SLESRC and PBSL. If a formal autopsy is not practicable, as is expected often to be the case based on prior experience conducting trials in similar settings, a verbal autopsy report will be completed and submitted to SLESRC, PBSL, and the WUSTL IRB. The verbal autopsy will be conduced in line with the World Health Organization guideline. If obtainable, the cause of death shall be classified according to the current ICD guideline.

The IDMC will receive information on all SAEs within 5 working days. The IDMC will independently review SAEs at least yearly. The IDMC will generate a report on SAEs for the PI, SLESRC, PBSL, and WUSTL IRB. Through its reviews of the study, the IDMC will determine whether cumulative data indicates the need to change the research design, to modify information presented to participants, or to terminate the project. Any action taken to suspend or terminate the project will be reported to the PI, SLESRC, PBSL, and WUSTL IRB.

###### 8.3.7 REPORTING EVENTS TO PARTICIPANTS

In the event that SAEs occur at a higher rate in the intervention group that in the control group, participants will be informed.

###### 8.3.8 EVENTS OF SPECIAL INTEREST

Not applicable

###### 8.3.9 REPORTING OF PREGNANCY

Evaluation for pregnancy by ultrasound will occur during screening for trial inclusion and is an inclusion criterion.

##### 8.4 UNANTICIPATED PROBLEMS

###### 8.4.1 DEFINITION OF UNANTICIPATED PROBLEMS (UP)

The Office for Human Research Protections considers unanticipated problems involving risks to participants or others to include, in general, any incident, experience, or outcome that meets **all** of the following criteria:

- Unexpected in terms of nature, severity, or frequency given (a) the research procedures that are described in the protocol-related documents, such as the Institutional Review Board (IRB)-approved research protocol and informed consent document; and (b) the characteristics of the participant population being studied;
- Related or possibly related to participation in the research (“possibly related” means there is a reasonable possibility that the incident, experience, or outcome may have been caused by the procedures involved in the research); and
- Suggests that the research places participants or others at a greater risk of harm (including physical, psychological, economic, or social harm) than was previously known or recognized.

###### 8.4.2 UNANTICIPATED PROBLEM REPORTING

When an unanticipated problem that is a SAE is suspected, the principal investigator must be informed within 24 hours and then must inform SLESRC, PBSL, and the WUSTL IRB within 24 hours. The UP report will include the following information:

- Protocol identifying information: protocol number, PI's name, and the IRB project number;
- A detailed description of the event, incident, experience, or outcome;
- An explanation of the basis for determining that the event, incident, experience, or outcome represents an UP;
- A description of any changes to the protocol or other corrective actions that have been taken or are proposed in response to the UP.

Any other UP will be reported to the IRB within 3 working days of the investigator becoming aware of the problem.

##### 8.4.3 REPORTING UNANTICIPATED PROBLEMS TO PARTICIPANTS

UPs will be reported to all participants who remain enrolled. Efforts will be made to contact participants who have completed the trial to inform them of UPs.

### 9 STATISTICAL CONSIDERATIONS

#### 9.1 STATISTICAL HYPOTHESES

All analyses will be compared under a superiority framework.

**Primary Efficacy Endpoints:**

- We hypothesize that adding 500mg DHA, 500mg EPA, and 550mg choline to a supplementary food provided to malnourished pregnant Sierra Leonean women with enrollment gestational age ≤ 30 weeks will prolong gestation (days) compared with provision of a similar supplementary food that does not contain DHA, EPA, or choline.
- We hypothesize that adding 500mg DHA, 500mg EPA, and 550mg choline to a supplementary food provided to malnourished pregnant Sierra Leonean women will improve offspring cognitive development at 9 months of age (MDAT global z-score) when compared with provision of a similar supplementary food that does not contain DHA, EPA, or choline.
- Among women enrolled in the above trial who develop ante- or post-natal depression, we hypothesize that providing a novel cognitive behavioral therapy (CBT) will improve depression (PHQ-9 score) compared with no CBT.

**Secondary Efficacy Endpoints**

- We hypothesize that adding 500mg DHA, 500mg EPA, and 550mg choline to a supplementary food provided to malnourished pregnant Sierra Leonean women with enrollment GA ≤ 30 weeks will:
  - Reduce early preterm birth (<34 weeks)
  - Increase birth length and weight
  - Reduce low birth weight (<2.5 kg)
  - Reduce preterm birth (<37 weeks)
  - Reduce neonatal mortality (live birth and subsequent death < 28 days from delivery)
- We hypothesize that adding 500mg DHA, 500mg EPA, and 550mg choline to a supplementary food provided to malnourished pregnant Sierra Leonean women will:
  - Improve MDAT sub-domain (gross motor, fine motor, language, social) z-scores
  - Prolong gestation
  - Reduce depressive symptoms (PHQ-9) prior to delivery
  - Increase maternal and cord blood RBC-DHA levels

#### 9.2 SAMPLE SIZE DETERMINATION

Trial sample size determination was done in reference to both primary outcomes, duration of gestation in days and MDAT global-z-score. The goal was to achieve at least 80% power for both primary outcomes. A simulation-based power analysis approach was undertaken. All analyses were done using a two-sided α of 0.05.

**Gestational Duration**

The population for analysis for this outcome variable will be women with enrollment GA ≤ 30 weeks.

A preliminary goal of powering that study to detect a difference gestational duration of 3 days was made based on a secondary analysis of the 2021 MamaDutasi trial[6] performed by our group amongst the same population, which showed a 3-day increase in gestational duration reduced the odds of neonatal death by 67% among preterm births (results not published). Results from this trial were also used to estimate the mean (for the control group) and SD (both groups) of gestational duration for preliminary power analysis.

In the prior trial, fundal heights were used to estimate gestational duration, with estimated gestational duration (days) = (final measured fundal height (cm) x 7) + time (days) between measurement and delivery date. In malnourished women, it is possible that this method underestimated gestational duration due to slower intrauterine growth and also might have affected the skew (e.g., if IUGR babies were more likely to be born preterm). However, the SD (16.5 days) closely resembles those commonly reported in other trials of DHA supplementation where GA was an outcome. Simulations were done to assess potential impact of different distributions, as detailed below.

Gestational duration has a left skew. The severity of skew can vary by setting. There are limited data in Sierra Leone on the distribution of gestational duration, and none are high-quality due to lack of ultrasound dating and high frequency of late presentations making LMP unreliable. We opted to use our 2021 trial data for this purpose. First, the 2021 gestational duration data were compared between intervention and control groups of that trial to assess whether there might be issues using parametric tests such as a t-test for this analysis. Doing so revealed that residuals violated normality (particularly at the left tail) and that there was heteroscedasticity. This prompted further investigation as to whether parametric or non-parametric tests would be better suited to evaluate this outcome variable. As a next step, the distribution of gestational duration from the 2021 trial was abstracted by day of gestational duration (e.g., 0.08% of deliveries occurred at 24 + 5 days... 0.24% of deliveries occurred at 31 + 3 days, 0.15% occurred at 31 + 4 days... 1.98% occurred at 37 + 6 days, and so on). 10,000 simulated datasets with 700 rows (participants) were generated from this distribution (group 1), by sampling with replacement. 3 days were added to the distribution (i.e., distribution shifted rightward by 3 days), and 10,000 further simulated datasets with 700 rows were generated (group 2), again by sampling with replacement. These simulated datasets were merged, and a t-test was used to compare group 1 vs. group 2. Again, non-normality of residuals and heteroscedasticity were identified. Thus, the decision was made to plan to use non-parametric tests to compare groups.

It is known that second trimester and third trimester gestational age estimations by last menstrual period (LMP) are unreliable, while GA estimations done by ultrasound with standard biometrics in the second and third trimesters have measurement error. Judging by the 2021 trial results, we expect the median (IQR) of enrollment GA to be approximately 24 (18-28). Using ultrasound dating is likely to shift this leftward to some extent (perhaps 2-3 weeks), as we expect growth restriction to impact fundal height more than US dating. In addition, attempts will be made to recruit at earlier GA in this trial, though it is not clear if this will be possible, in part because there are no high-quality data on when undernutrition develops during pregnancy. It may be that some pregnant women develop undernutrition primarily during second/third trimesters and thus would not be eligible for enrollment until that time. Thus, it was assumed that most women will be enrolled during the second and third trimesters. Using ultrasound, GA estimation in second trimester is considered to add +/- 14 days of error to the estimate (2SD = 14 days). This error can be added to the outcome measure using simulation-based power analysis.

Ten thousand simulations were undertaken wherein control and intervention groups were simulated using the aforementioned distribution (control) or the aforementioned distribution + 3 days (intervention). Random errors with mean = 0 and SD = 7 days were then added to both groups' distributions. The Wilcoxon-Mann-Whitney test was used to compare groups across all simulations and the percent with p-values < 0.05 was determined. Across iterations, it was determined that 510 individuals per group were needed to achieve 80% power; the distribution is shown below. All power analyses were done using R.

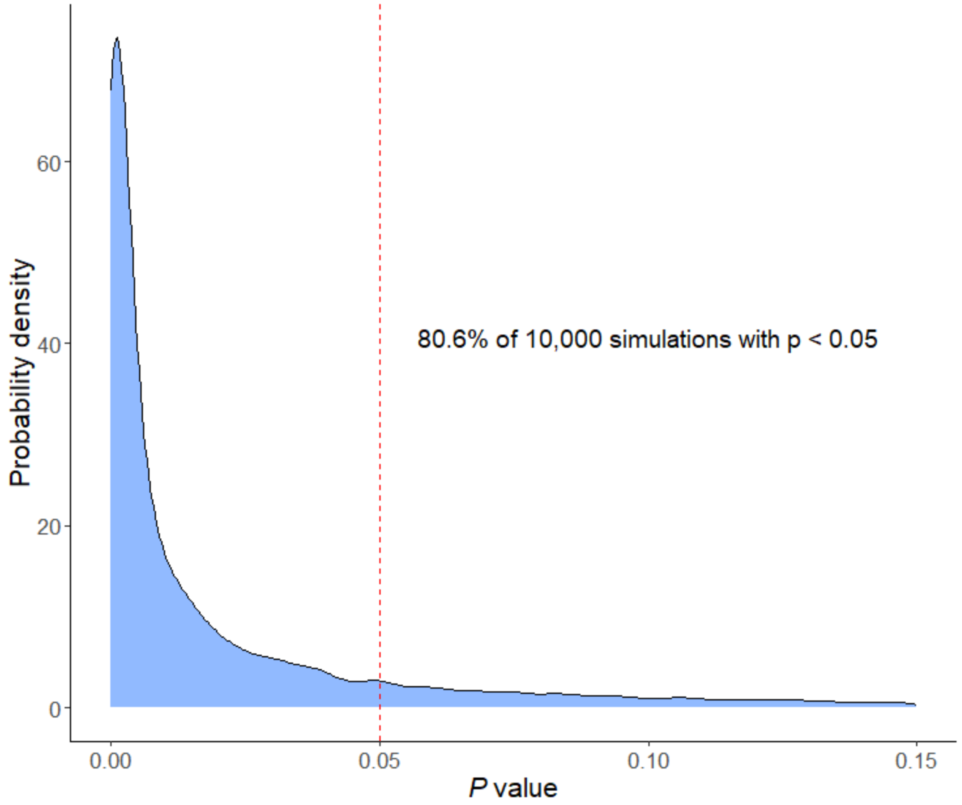

In the 2021 trial, 85% of participants remained in the trial (i.e., did not default) until delivery and had live singleton births. Assuming a similar 15% loss prior to primary outcome (gestational duration) determination, 510 participants per group for the primary outcome will require enrollment of 600 participants with enrollment GA ≤ 30 weeks per group, which will itself require 800 participants total per group. This estimation of 25% enrollment GA > 30 weeks is also drawn from the 2021 trial. In that study, 15% of participants had enrollment fundal height > 30 weeks. We expect fundal height to have underestimated GA by 2-3 weeks at fundal heights > 30 weeks due to intrauterine growth restriction. Evidence for this estimate comes from assessing birth weights across estimated gestational duration, which suggest a 2-3 week shift.

**Malawi Developmental Assessment Tool**
For the second primary outcome – MDAT at 9 months of age – 800 participants per group will provide 80% power at a two-sided α of 0.05 to detect a 0.2 z-score increase in global MDAT score among offspring, assuming 28% of participants/offspring will either default or not be alive at 9 months of age, leaving 576 per group. The assumptions underlying this power analysis were drawn from two prior studies from our group, the Improved PUFA trial and the 2021 MamDutasi trial. The target 0.2 z-score

increase is close to the 0.19 difference detected in the Improved PUFA trial, which tested adding DHA to therapeutic food for severely malnourished children. The 28% default/death estimation is based on the MamDutasi trial, in which 25% of participants/offspring either were lost to follow-up or had died by 6 months after delivery. We assume 3% more participants will default between 6 -> 9 months based on the rate per month seen in that study.

The power analysis was done two ways. The expected distribution of MDAT scores in these 9mo children whose mothers were malnourished is unknown. First, a normal distribution was assumed, with control group mean = 0 and SD = 1.2, intervention group mean = 0.2 and SD = 1.2. This SD was drawn from the Improved PUFA trial. It is possible that the history of severe acute malnutrition among the children in this study increased the SD above what would be expected in COGENT; however, all of the children in COGENT will have been born to malnourished mothers, which also may increase the SD of their scores. Using a t-test, 576 participants per group provided 80.7% power across 10,000 simulations.

Second, given the uncertainty regarding the distribution and potential for left skew (common for developmental tests, more likely among a disadvantaged population), an alternative approach was undertaken. The distribution of MDAT z-scores from the Improved PUFA trial was abstracted. For the control group, 10,000 simulated datasets of populations sampled with replacement from this distribution were generated. 5,000 such simulations are plotted below, in which each line represents a single sample and drawn along a scale from yellow to green. As displayed, the distributions can vary by 0.5 – 1 z-score.

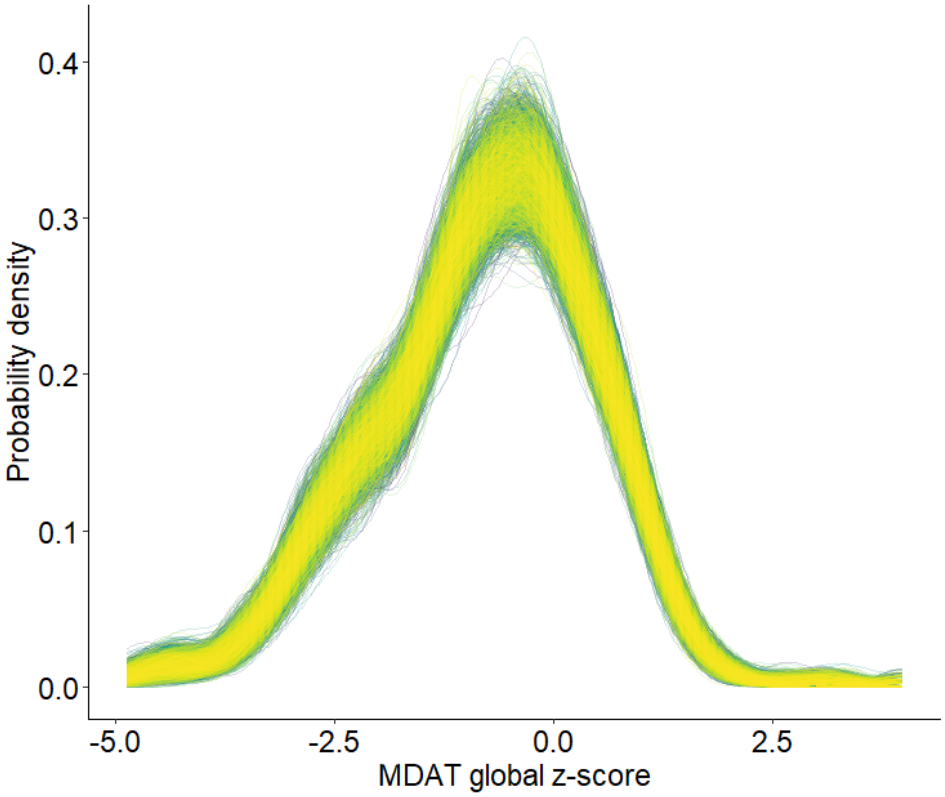

For the intervention group, 0.2 z-scores were added to the parent distribution and 10,000 simulated datasets were again generated, then merged with the control data. The Wilcox-Mann-Whitney test was used to compare all 10,000 simulated datasets and the power was 80%.

**PHQ-9 score**

The study was not powered specifically for the CBT vs. no CBT comparison, which will only apply to the subgroup of participants who develop depression (PHQ-9 ≥ 9) during the trial. For the CBT vs. no CBT primary outcome, 64 participants per group will provide >90% power to detect a decrease of 1.5 points on the PHQ-9 score at a two-sided α of 0.05. This power analysis was done via simulation. The baseline distribution of PHQ-9 scores among malnourished pregnant women in Sierra Leone with scores ≥ 9 is not known. 50,000 simulated distributions with varying medians and degrees of skewness and kurtosis were generated using the Dirichlet distribution and a random vector generator. A cumulative probability plot displaying 5,000 such simulated distributions each are shown below. The "parent" distribution from which the alternatives were simulated assumes a large proportion participants will have scores on the lower end of the PHQ-9 ≥ 9 spectrum, as is reflected in the plot. The random vector generator produces alternative distributions at degrees of similarity specified by the user. 5 different degrees of similarity were used, with 10,000 simulations per degree.

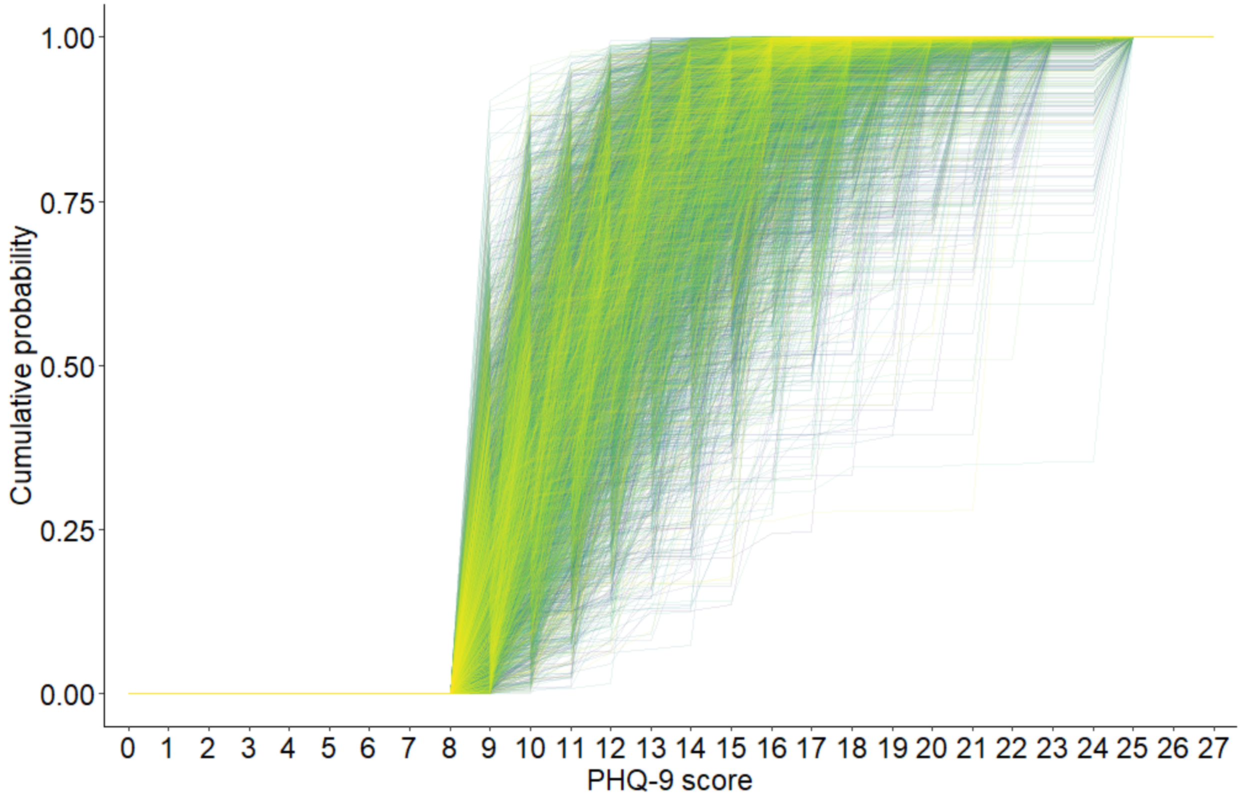

64 participants per group provided 90% power across the 50,000 simulations to detect a median difference between groups in PHQ-9 score of 1.7 (IQR 1.1, 2.2) using ordinal logistic regression, corresponding to an odds ratio for better PHQ-9 score of 3. Based on a pilot program undertaken in Pujehun District in Sierra Leone June 2022 – October 2022, we estimate that 10% of malnourished pregnant women will develop PHQ-9 scores ≥ 9. We expect 10-15% of participants to drop out during the prenatal period, and a further 10% to default in the postnatal period. Thus, assuming 20% of participants will not be assessable on average during the trial, 800 participants per group will provide sufficient sample size for this outcome.

#### 9.3 POPULATIONS FOR ANALYSES

Primary analyses will be done using two modified intention-to-treat (mITT) datasets, one for gestational duration, and the other for MDAT global z-scores.

For both mITT datasets, the modifications are as follows: (1) participants who are found to have an exclusion criterion after enrollment/randomization will be excluded when it is identified, (2) participants who are lost to follow-up or experience stillbirth or miscarriage will be excluded from analysis, and (3) mothers who pass away prior to delivery will be excluded from analysis.

For MDAT, participants who do not undergo MDAT testing (lost to follow-up, infant death) will be excluded from analysis.

For gestational duration, this will only be assessed among women with enrollment GA ≤ 30 weeks. This decision was made as a compromise between arguments in favor of more restrictive enrollment criteria:

- preceding data from studies showing DHA benefit generally only included women with enrollment GA < 20 weeks
- the mechanisms underlying pre-term birth might begin earlier in pregnancy
- there is a hypothesis that most benefit is derived from shifting early pre-term births right-ward

and those in favor of more broad criteria:

- many women in LMICs present later in pregnancy, and thus results of a trial restricting enrollment to < 20 weeks would likely not apply to many of these women
- for some women, undernutrition does develop during the second and third trimesters of pregnancy, and it would be valuable to know if this intervention might help these women as well
- undernutrition is a state of total-body depletion, and thus it is possible that DHA supplementation even later in pregnancy might benefit these women and their children

Thus, ≤ 30 weeks was chosen as a compromise cut-off for gestational duration co-primary outcome.

All participants will be included in safety analyses.

In a per-protocol analysis, only participants who missed 0 study visits will be included.

Sub-group analyses will be done on the basis of baseline RBC-DHA content quartiles, baseline MUAC, enrollment gestational age, parity, and maternal age.

#### 9.4 STATISTICAL ANALYSES

##### 9.4.1 GENERAL APPROACH

Categorical variables will be summarized as n (%). Continuous variables will first be plotted using probability density plots. If their distribution approximates normality, they will be summarized as mean (SD); otherwise, they will be summarized as median (IQR). Several key variables, such as gestational duration and MDAT, may be summarized using both. *P* values will be considered statistically significant when < 0.05. 95% CIs will be estimated for all between-group comparisons, including differences, ORs, and RRs. Neither *P* values nor 95% CIs will be adjusted for multiple comparisons.

##### 9.4.2 ANALYSIS OF THE PRIMARY EFFICACY ENDPOINT(S)

Analyses will be performed on the modified intention-to-treat populations outlined **in section 9.3**. There will be no adjustment for multiplicity. The factorial components can be considered separate trials; it is expected that EPA and CBT could be additive in their effects but are not expected to be multiplicative. For co-primary outcomes gestational duration and MDAT in the M-RUSF+ vs. M-RUSF comparison, in this trial of a low-risk intervention in which outcomes may be correlated, it is felt to be more important to maintain study power than guard against inflating type I error.

**Gestational duration**
This will be calculated by adding the enrollment estimated gestational age (days) to the number of days between enrollment and date of delivery. Unadjusted analysis will be done using the Wilcoxon-Mann-Whitney test. The median of differences (likely estimated using the Hodges-Lehmann estimator) with a 95% CI will be presented. Adjusted analysis will be done using ordinal logistic regression. The proportional odds assumption will be assessed for treatment group; if deemed violated, an alternative will be chosen, such as partial proportional odds ordinal logistic regression. If proportional odds are violated for a covariate, this will not change the analytic plan, as such a violation has been shown to be unlikely to impact effect estimates. Covariates will be maternal age and number of prior pregnancies, which were chosen after evaluating possible predictors of gestational duration in malnourished pregnant women in Sierra Leone from our 2021 trial. OR (95% CI) and difference in model derived medians with bootstrapped 95% CI will be presented.

**Malawi Developmental Assessment Tool**
This will be calculated by the tablet program as an age-adjusted global z-score. Residual normality and homoscedasticity will be evaluated prior to use of linear regression. If assumptions are met, linear regression will be used to compare groups, with difference (95% CI) reported. If assumptions are not met, Wilcoxon-Mann-Whitney test will be done, with median of differences and 95% CI reported. This procedure is necessary because we do not know / cannot predict how MDAT modeling errors will distribute in 9 mo children born to Sierra Leonean women who were malnourished during pregnancy. Similarly, it is unclear which variables might be valuable for adjustment. As a result, adjusted analyses will be considered exploratory, with possibilities including gestational age at enrollment, gestational weight/length, sex, maternal education level, maternal HIV status, and number of children sleeping in home.

**Patient Health Questionnaire-9**
Scores will be calculated by adding up the points across the 9-element test. This score is ordinal in nature and will be collected in a repeated manner bi-weekly during and for 4 weeks after treatment (baseline + ~5 follow-up tests). Longitudinal ordinal logistic regression will be used to model CBT vs. no CBT effect on PHQ-9 over time. Participant ID will be a random effect, including slope. It is anticipated that treatment x time interaction term will be used to compare groups. The output will be OR (95% CI).

##### 9.4.3 ANALYSIS OF THE SECONDARY ENDPOINT(S)

These pre-planned secondary analyses will not depend on results of primary outcome analyses. Analyses will be performed on the modified intention-to-treat population. There will be no adjustment for multiplicity.

For the M-RUSF+ vs. M-RUSF comparison:

**Early preterm birth**
ePTB will be defined as duration of gestation (enrollment GA estimate + time between enrollment and date of delivery) < 34 weeks. It is a binary outcome. Logistic regression will be used to compare groups, with OR (95% CI) estimated.

**Birth weight and length**
These will be measured directly in duplicate with measurements averaged, often within 1 day of delivery and almost always within 2 days of delivery. Following expected confirmation of residual normality and homoscedasticity, values will be compared between treatment groups using linear regression with differences (95% CIs) estimated.

**Low birth weight**
PBW will be defined as birth weight < 2.5 kg. It is a binary outcome. Logistic regression will be used to compare groups, with OR (95% CI) estimated.

**MDAT sub-domain z-scores**
The same procedure will be used as described above for the MDAT global z-score primary efficacy outcome.

**Neonatal mortality**
Neonatal mortality will be defined as live birth with subsequent death < 28 days from date of birth. It is a binary outcome. Time-to-event analysis will be used to compare groups, with censoring for default during follow-up. The proportional hazards assumption will be assessed prior to use of Cox proportional hazards regression. HR (95% CI) will be estimated.

**Preterm birth**
PTB will be defined as duration of gestation (enrollment GA estimate + time between enrollment and date of delivery) < 37 weeks. It is a binary outcome. Logistic regression will be used to compare groups, with OR (95% CI) estimated.

**PHQ-9 scores**
PHQ-9 scores will be tabulated by summing points on the 9-item scale. This is a longitudinal ordinal outcome. Longitudinal ordinal logistic regression will be used to compare groups, with random effect for participant intercept and slope. Treatment x time interaction will be the parameter of interest. OR with 95% CI will be estimated.

**RBC-DHA content**
Expressed as % of fatty acids. This variable is expected to have non-normal residuals and heteroscedasticity. Analysis will be done using the Wilcoxon-Mann-Whitney test. Median of differences and 95% CI will be estimated.

For M-RUSF+ vs. M-RUSF and CBT vs. no CBT comparisons:

**PHQ-9 < 9**
Binary outcome for "resolution of depression". Logistic regression will be used to compare groups. Analyses will be adjusted for baseline PHQ-9 (M-RUSF+ vs. M-RUSF) or PHQ-9 at time of depression diagnosis (CBT vs. no CBT). ORs with 95% CIs will be estimated.

##### 9.4.4 SAFETY ANALYSES

The primary safety endpoints will be maternal death, infant death, miscarriage, stillbirth, postnatal delivery (>42 weeks), maternal prepartum hospitalization, and peripartum complication requiring transfer to facility with higher level of care. These will likely be presented in a table using summary statistics and compared between groups using logistic regression. Gastrointestinal adverse events (vomiting, diarrhea) will also be reported.

##### 9.4.5 BASELINE DESCRIPTIVE STATISTICS

Baseline descriptive statistics will be presented as per the procedure outlined in **section 9.4.1.** There will be no statistical comparison between groups.

##### 9.4.6 PLANNED INTERIM ANALYSES

Not applicable

##### 9.4.7 SUB-GROUP ANALYSES

Sub-group analyses for primary and secondary endpoints in the M-RUSF+ vs. M-RUSF comparison will be done on the basis of baseline RBC-DHA content, baseline MUAC, enrollment gestational age, parity, and maternal age. We will likely model "sub-groups" as interaction terms between study group and variable of interest with the variable of interest taken as a continuous variable, with spline terms included if the relationship appears non-linear. Baseline RBC-DHA content was chosen because of prior RCT results suggesting greater benefit of prenatal DHA supplementation among mothers with lower baseline DHA status. Baseline MUAC was chosen as a surrogate for degree of undernutrition; it is possible the intervention might have differential impacts on this basis. Enrollment gestational age was chosen because most prior trials showing benefit of DHA enrolled more stringently, e.g. GA < 20 weeks, and it is possible those who receive supplementation longer might derive more benefit. Maternal age was chosen on the basis of a secondary analysis of the prior Sierra Leone maternal undernutrition study by our group, wherein offspring of adolescent mothers were found to derive less benefit from the combined intervention of food + anti-infective measures than adult mothers.[34] Parity was chosen because it predicts gestational duration.

Subgroups analysis for PHQ-9 score for the CBT vs. no CBT comparison will be done for ante- vs. postpartum depression and baseline PHQ-9 score.

###### 9.4.8 TABULATION OF INDIVIDUAL PARTICIPANT DATA

Individual participant data will not be provided in the study report.

###### 9.4.9 EXPLORATORY ANALYSES

We will explore whether M-RUSF+ might reduce pre-term birth (gestational duration < 37 weeks), the incidence of depression (PHQ-9 ≥ 9), increase placental weight, increase birth chest and head circumference, and alter breast milk human milk oligosaccharides. We will explore whether CBT vs. no CBT might prolong gestation and improve infant cognitive development.

If M-RUSF+ prolongs gestation and improves MDAT global z-scores, we will perform a mediation analysis with gestational duration as the mediating variable.

### 10 SUPPORTING DOCUMENTATION AND OPERATIONAL CONSIDERATIONS

#### 10.1 REGULATORY, ETHICAL, AND STUDY OVERSIGHT CONSIDERATIONS

##### 10.1.1 INFORMED CONSENT PROCESS

###### 10.1.1.1 CONSENT/ASSENT AND OTHER INFORMATIONAL DOCUMENTS PROVIDED TO PARTICIPANTS

Consent forms describing in detail the study intervention, study procedures, and risks are given to the participant and written (or thumb print) documentation of informed consent is required prior to administering study intervention.  The following consent materials are submitted with this protocol:

M-RUSF+ vs. M-RUSF study consent
M-RUSF+ vs. M-RUSF study assent
CBT vs. no CBT study consent
CBT vs. no CBT study assent

###### 10.1.1.2 CONSENT PROCEDURES AND DOCUMENTATION

Head chiefs for all catchment areas will be informed of the study and their consent will be obtained prior to trial initiation. This process has been part of the clinical trials routine for the PI's research group in Malawi and Sierra Leone for two decades and has been successful.

All consent processes will be performed by Sierra Leonean research nurses in participant native language (e.g., Mende, Krio, etc.). Based on prior experience, it is expected that most participants will not be literate. Thus, much of the consent process is expected to be verbal in nature, with thumbprint consent documentation in the event of participant consent or assent. Because so much of the consent process will rely on our research nurses verbalizing study procedures, potential benefits, risks etc., these nurses will undergo extensive education and training around the trial and consent processes prior to study initiation. This process is essential for the trial to reach as many of the most vulnerable Sierra Leonean women as possible.

Informed consent is a process that is initiated prior to the individual's agreeing to participate in the study and continues throughout the individual's study participation. Consent forms will be SLESRC and WUSTL IRB-approved and the participant will be asked to read and review the document if they are literate; otherwise, Sierra Leonean research nurses will read and review the document for participants in their native language. The Sierra Leonean research nurses will explain the research study to the participant and answer any questions that may arise. A verbal explanation will be provided in terms suited to the participant's comprehension of the purposes, procedures, and potential risks of the study and of their rights as research participants. Participants will have the opportunity to carefully review the written consent form and/or ask questions prior to signing. The participants will have the opportunity to discuss the study with their family or surrogates or think about it prior to agreeing to participate. The participant will sign or, if illiterate, apply a thumbprint to the informed consent document prior to joining the study. Participants will be informed that participation is voluntary and that they may withdraw from the study at any time, without prejudice. The informed consent process will be conducted and documented in the source document (including the date), and the form signed, before the participant undergoes any study-specific procedures. The rights and welfare of the participants will be protected by emphasizing to them that the quality of their medical care will not be adversely affected if they decline to participate in this study.

In the event of enrolling a participant under 16 years of age, parental/guardian consent will be required, with participant assent. The same procedures as described above will be undertaken.

##### 10.1.2 STUDY DISCONTINUATION AND CLOSURE

This study may be temporarily suspended or prematurely terminated if there is sufficient reasonable cause. If the study is prematurely terminated or suspended, the PI will promptly inform study participants, the IRBs, and sponsor and will provide the reason(s) for the termination or suspension. Study participants will be contacted, as applicable, and be informed of changes to the schedule of study visits.

Circumstances that may warrant termination or suspension include, but are not limited to:

- Determination of unexpected, significant, or unacceptable risk to participants
- Insufficient compliance to protocol requirements
- Data that are not sufficiently complete and/or evaluable

Study may resume once concerns about safety, protocol compliance, and data quality are addressed, and satisfy the PI and IRB.

##### 10.1.3 CONFIDENTIALITY AND PRIVACY

Participant confidentiality and privacy are strictly held in trust by the participating investigators and their staff. This confidentiality is extended to cover testing of biological samples and genetic tests in addition to the clinical information relating to participants. Therefore, study data and all other information generated will be held in strict confidence. No information concerning the study or the data will be released to any unauthorized third party without PI approval.

All research activities will be conducted in as private a setting as possible. Because the study will largely take place at government-run antenatal clinics, there will be limitations as to the degree of privacy possible. However, for sensitive activities such as gestational age ultrasounds and cognitive behavioral therapy, privacy will be maintained by using a separate room or facility.

Study participant contact information will be securely stored at the two research bases of operation in Pujehun and Bo Districts in Sierra Leone for internal use during the study. At the end of the study, all records will continue to be kept in a secure location for as long a period as dictated by the reviewing IRB.

Study participant research data, which is for purposes of statistical analysis and scientific reporting, will be transmitted to and stored on a WUSTL-based secure cloud computing site, Box. An exception is ultrasound videos, which will initially be stored using the secure Butterfly App until service is available to upload the videos to WUSTL Box, after which point they will be purged from the Butterfly App. Such stored data will not include the participant’s contact or identifying information. Rather, individual participants and their research data will be identified by a unique study identification number. The study data entry and study management systems used by research staff will be secured and password protected. At the end of the study, all study databases will be de-identified and archived securely at WUSTL.

##### 10.1.4 FUTURE USE OF STORED SPECIMENS AND DATA

Data collected for this study will be analyzed and stored at WUSTL in the PI’s secure cloud database. After the study is completed, the de-identified, archived data will be transmitted to and stored at WUSTL, for use by other researchers including those outside of the study once approval has been obtained by the PI.

With the participant’s approval and as approved by local IRBs, de-identified biological samples will be stored at WUSTL in the PI’s laboratory freezers. These samples could be used to study various physiologic and pathologic processes, including but not limited to preterm labor, neonatal mortality, lipid metabolism and its genetic implications, DHA trafficking, metabolomics, infant growth, and cognitive development. The potential for future use will be explicit in the trial consent forms. The WUSTL repository will also have a code-link that will allow linking the biological specimens with the phenotypic data from each participant, maintaining the blinding of the identity of the participant. This potential use will be made clear in the consent.

During the conduct of the study, an individual participant can choose to withdraw consent to have biological specimens stored for future research. However, withdrawal of consent with regard to bio-sample storage may not be possible after the study is completed, as there is unlikely to be contact with study participants after their involvement has finished.

##### 10.1.5 KEY ROLES, STUDY TEAM, AND STUDY GOVERNANCE

Principal Investigator: Mark Manary, MD, Washington University in St. Louis
Co-PI: Kevin Stephenson, MD, Washington University in St. Louis
Co-Investigator: D. Taylor Hendrixson, MD, University of Washington
Study Monitor: Donna Wegner, Washington University in St. Louis
National PI, Aminata Koroma MS. Director of Nutrition, Ministry of Health & Sanitation of Sierra Leone
Pujehun District Collaborator, Dr. Amara Stevens Ngegbai MD
Ibrahim Affie Bangura, Director of Finances and Human Resources
Joshua Abioseh Duncan, Coordinator for Cognitive Behavioral Therapy, Mental Health Coalition of Sierra Leone
Julius Butcher, Technical Supervisor for Clinical Staff, Project Peanut Butter
Chief of Food Production, Tabita Kamara PhD, Project Peanut Butter
Research Associate TBA, Washington University in St. Louis
Nurse-midwives (4) fluent in local language Mende, having completed nursing school and midwife program and with 5 years' experience, Project Peanut Butter
Counselors, Project Peanut Butter
Cognitive development assessment team, Project Peanut Butter
Drivers, Project Peanut Butter

##### 10.1.6 SAFETY OVERSIGHT

The data and safety monitoring plan will involve a chain of reporting of AEs and SAEs within predetermined time windows as indicated in section 8.3. All research staff will be empowered to report safety concerns to the Research Associate (RA).

##### 10.1.7 CLINICAL MONITORING

Clinical site monitoring is conducted to ensure that the rights and well-being of trial participants are protected, that the reported trial data are accurate, complete, and verifiable, and that the conduct of the trial is in compliance with the currently approved protocol/amendment(s), with International Conference on Harmonisation Good Clinical Practice (ICH GCP), and with applicable regulatory requirement(s).

The foundational component of study monitoring will be the responsibility of all research staff who interact with participants, provide clinical care, and collect data. The nurse-midwives, RA, and visiting MDs will ensure professional and appropriate clinical care for participants each study day. There will be frequent training for consenting, ultrasonography, anthropometric measurements, and food disbursement to ensure integrity of study procedures. Daily checks of all data cards prior to participants leaving the clinic will be done to evaluate for missing or illogical data. Participants will be asked to bring back their empty sachets each visit to assess for correct food disbursement. Medication provision will be documented with dates on study cards. Supply lists will be kept updated daily to ensure there are no stock-outs.

In addition, the PI Mark Manary, Kevin Stephenson, and Taylor Hendrixson will provide on-site monitoring at least every 2 months throughout the study. This will involve visit clinics each day with the research team, overseeing each aspect of care and data collection, and evaluation of data entry.

The database will be checked each week remotely from St. Louis for missing data and illogical entries. Double entry from primary data cards will be done and checked against the initial entry for errors.

##### 10.1.8 QUALITY ASSURANCE AND QUALITY CONTROL

As stated in section 10.1.7, the most important aspect of quality assurance and control will be the daily work of the research staff in the field in Sierra Leone. Protocols have been developed for each technical procedure, including ultrasonography, blood spot collection, cord blood collection, and placental weight measurement. All relevant individuals will undergo training prior to trial initiation and intermittently during the trial at least q6mo or more frequently if issues arise. All data collected will be checked daily prior to discharging a participant from clinic to assess for missingness and implausibility. There will be checks on accuracy of study food disbursement and timeliness of medication administration by comparison of expectations based on scheduling with documented provision. There will be assessments as to the quality of CBT with random visits by the clinical coordinator overseeing the counselors. Data cards will be scanned and transmitted to the secure cloud database weekly alongside the updated database for review for completeness and plausibility. Any missing data or data anomalies will be communicated to the sites for clarification/resolution.

##### 10.1.9 DATA HANDLING AND RECORD KEEPING

###### 10.1.9.1 DATA COLLECTION AND MANAGEMENT RESPONSIBILITIES

Data collection is the responsibility of the clinical trial staff at the site under the supervision of the Research Associate. The RA is responsible for ensuring the accuracy, completeness, legibility, and timeliness of the data reported. The RA will have oversight in the form of remote monitoring using the cloud repository with weekly assessments of all data collected as well as in-person oversight with at least bi-monthly visits from the PI or designees.

Standardized hardcopy study cards will be used for recording data for each participant enrolled in the study. After clinic, these data will be entered daily into a secured electronic database at the research centers in Sierra Leone, followed by upload to the WUSTL-based secure cloud repository. Hardcopy study cards will also be scanned and uploaded to the same repository weekly. All data will be double-entered by different individuals. Discrepancies will be assessed by review of the primary data card.

Ultrasound videos will initially be stored on the Butterfly App prior to upload to the secure cloud-based repository once WiFi is accessible, after which point they will be deleted from the Butterfly App.

###### 10.1.9.2 STUDY RECORDS RETENTION

Study documents should be retained for a minimum of 6 years after the study has completed, in line with the WUSTL IRB policy.

###### 10.1.10 PROTOCOL DEVIATIONS

A protocol deviation is any noncompliance with the clinical trial protocol. The noncompliance may be either on the part of the participant, the investigator, or the study site staff. As a result of deviations, corrective actions are to be developed by the site and implemented promptly.

It is the responsibility of research staff to use continuous vigilance to identify deviations. It is the responsibility of the lead research assistants in Sierra Leone to report deviations within 1 working day of identification of the protocol deviation. All deviations must be addressed in study source documents and reported to the Principal Investigator. Protocol deviations must be sent to the reviewing Institutional Review Board (IRB) per their policies. All research team members are responsible for knowing and adhering to the reviewing IRB requirements.

###### 10.1.11 PUBLICATION AND DATA SHARING POLICY

The PI will determine authorship on primary and secondary publications of data derived from this trial and published by the PI's research group. De-identified data will be shared with PI approval beginning 6 months after primary trial publication.

###### 10.1.12 CONFLICT OF INTEREST POLICY

The funding agency will play no role in the design, implementation, analysis, or decision to publish results of this trial. Persons with actual conflicts of interest will not be allowed to participate in the trial in any manner. Furthermore, persons who have a perceived conflict of interest will be required to have such conflicts managed in a way that is appropriate to their participation in the design and conduct of this trial.

##### 10.2 ADDITIONAL CONSIDERATIONS

Not applicable

#### 10.3 ABBREVIATIONS

| | |
|---|---|
| AE | Adverse event |
| BMI | Body-mass index |
| CBT | Cognitive behavioral therapy |
| CFR | Code of Federal Regulations |
| CHW | Community health worker |
| FAMLI | Fetal Age Machine Learning Initiative |
| GA | Gestational age |
| ICH GCP | International Conference on Harmonization Good Clinical Practice |
| IRB | Institutional Review Board |
| LAZ | Length-for-age z-score |
| MDAT | Malawi Developmental Assessment Tool |
| M-RUSF | Maternal ready-to-use supplementary food |
| M-RUSF+ | Maternal ready-to-use supplementary food+ |
| mITT | Modified intention-to-treat |
| MUAC | Mid-upper arm circumference |
| PHQ-9 | Patient Health Questionnaire-9 |
| PPB | Project Peanut Butter |
| RA | Research Associate |
| RUSF | Ready-to-use supplementary food |
| RUTF | Ready-to-use therapeutic food |
| SAE | Serious adverse event |
| SLESRC | Sierra Leone Ethics Review Committee |
| UP | Unanticipated problem |
| US | United States of America |
| WLZ | Weight-for-length z-score |
| WUSTL | Washington University in St. Louis |

#### 10.4 PROTOCOL AMENDMENT HISTORY

| Version | Date | Description of Change | Brief Rationale |
|---|---|---|---|

30. *Dietary Reference Intakes for Vitamin A, Vitamin K, Arsenic, Boron, Chromium, Copper, Iodine, Iron, Manganese, Molybdenum, Nickel, Silicon, Vanadium, and Zinc*. 2001.

31. *Dietary Reference Intakes for Vitamin C, Vitamin E, Selenium, and Carotenoids*. 2000.

32. James G, Stephenson K, Callaghan-Gillespie M, et al. Docosahexaenoic Acid Stability in Ready-to-Use Therapeutic Food. *Foods*. 2023-01-09 2023;12(2):308. doi:10.3390/foods12020308

33. Pokaprakarn T, Prieto JC, Price JT, et al. AI Estimation of Gestational Age from Blind Ultrasound Sweeps in Low-Resource Settings. *NEJM Evidence*. 2022-04-26 2022;1(5)doi:10.1056/evidoa2100058

34. Koroma AS, Ellie M, Bangura K, et al. Supplementary feeding and infection control in pregnant adolescents—A secondary analysis of a randomized trial among malnourished women in Sierra Leone. *Maternal & Child Nutrition*. 2023-01-01 2023;19(1)doi:10.1111/mcn.13456

Appendix 2. Comparison of selected baseline participant characteristics between participants with known primary outcome and those with unknown primary outcome[1]

| | Known primary outcome | | Unknown primary outcome | |
|---|---|---|---|---|
| **Characteristic** | **CBT (n = 75)** | **Control (n = 65)** | **CBT (n = 4)** | **Control (n = 9)** |
| Age, years, median (IQR)[2] | 19 (18, 22) | 19 (18, 23) | 19 (18, 27) | 21 (18, 23) |
| MUAC at enrollment, cm, median (IQR) | 22.7 (22.0, 23.0) | 22.4 (21.4, 22.9) | 22.0 (20.9, 22.2) | 22.0 (20.4, 22.6) |
| Weeks in supplementary feeding trial before enrollment in this trial, median (IQR)[3] | 4 (0, 10) | 4 (0, 10) | 11 (4, 31) | 0 (0, 4) |
| 0 | 36 (48) | 28 (43) | 1 (25) | 5 (56) |
| ≤ 4 | 14 (19) | 9 (14) | 0 (0) | 2 (22) |
| > 4 | 25 (33) | 28 (43) | 3 (75) | 2 (22) |
| Pregnancy status at enrollment | | | | |
| Antenatal | 68 (91) | 57 (88) | 2 (50) | 7 (78) |
| Gestational age at enrollment, weeks, median (IQR)[4] | 22.5 (19.0, 28.2) | 20.9 (17.2, 26.1) | 20.8 (8.0, 33.6) | 24.4 (21.0, 27.3) |
| Postnatal | 7 (9) | 8 (12) | 2 (50) | 2 (22) |
| Weeks since delivery at enrollment | 13.2 (12.3, 13.9) | 6.6 (6.2, 10.0) | 26.7 (13.7, 39.6) | 5.7 (5.4, 6.0) |
| Mother's education level | | | | |
| None | 13 (17) | 18 (28) | 1 (25) | 3 (33) |
| Primary | 6 (8) | 10 (15) | 1 (25) | 1 (11) |
| Secondary or greater | 56 (75) | 37 (57) | 2 (50) | 5 (56) |
| Currently in school[5] | 30 (40) | 16 (25) | 2 (50) | 0 (0) |
| History of miscarriage or stillbirth | 2 (3) | 6 (9) | 0 (0) | 1 (11) |
| Number of adults in household, median (IQR) | 4 (3, 5) | 4 (3, 5) | 2 (2, 4) | 4 (2, 6) |
| Two or more children live in household | 55 (73) | 52 (80) | 3 (75) | 6 (67) |
| Father lives in household | 51 (68) | 42 (65) | 3 (75) | 5 (56) |
| Household roof made of thatch | 8 (11) | 8 (12) | 2 (50) | 0 (0) |
| Pit latrine used for stool disposal | 63 (84) | 53 (82) | 4 (100) | 8 (89) |
| Household has electricity | 7 (9) | 11 (17) | 0 (0) | 0 (0) |
| Household Food Insecurity Access Scale score (past 4 weeks) | | | | |
| Food secure | 2 (3) | 7 (11) | 0 (0) | 1 (11) |
| Mildly food insecure | 0 (0) | 1 (1) | 0 (0) | 0 (0) |
| Moderately food insecure | 31 (41) | 18 (28) | 1 (25) | 3 (33) |
| Severely food insecure | 42 (56) | 39 (60) | 3 (75) | 5 (56) |
| aPHQ-9 score at time of enrollment, median (IQR) | 11 (10, 12) | 11 (9, 12) | 12 (10, 13) | 10 (9, 11) |
| Score of 9 | 10 (13) | 17 (26) | 1 (25) | 4 (44) |
| Score 10-12 | 50 (67) | 39 (60) | 2 (50) | 5 (56) |
| Score ≥ 13 | 15 (20) | 9 (14) | 1 (25) | 0 (0) |

[1] Values are n (%) unless otherwise indicated
[2] n = 2 missing from CBT and n = 1 missing from control, both with known primary outcomes
[3] CBT trial participants are a subset of a larger supplementary feeding study, details of which can be found in the Appendix
[4] n = 2 missing from control due to miscarriage prior to gestational age estimation being possible, both with known primary outcomes
[5] n = 1 missing from control, with unknown primary outcome

Appendix 3. Best-case sensitivity analysis for simulated ITT intervention effects on primary and secondary outcomes[1]

| | CBT | Control | Comparison[2] (95% CI) | *P* value[3] |
|---|---|---|---|---|
| Primary outcome | n = 79 | n = 74 | | |
| Endline aPHQ-9 | 2 (1, 4) | 5 (2, 9) | -3 (-4, -2) | <0.001 |
| Secondary outcomes | n = 79 | n = 74 | | |
| Change in aPHQ-9, endline - baseline | -9 (-10, -7) | -5 (-8, -2) | -4 (-5, -3) | <0.001 |
| Reduction in aPHQ-9 from baseline, n (%) | | | | |
| ≤ 3 | 3 (4) | 29 (39) | 0.06 (0.01, 0.18) | <0.001 |
| > 3 | 76 (96) | 45 (61) | 16.33 (5.41, 70.93) | <0.001 |
| > 5 | 70 (89) | 35 (47) | 8.67 (3.92, 20.95) | <0.001 |
| > 9 | 30 (38) | 4 (5) | 10.71 (3.92, 37.80) | <0.001 |
| > 50% | 72 (91) | 38 (51) | 9.74 (4.17, 25.80) | <0.001 |
| Endline aPHQ-9 < 5 | 63 (80) | 31 (42) | 5.46 (2.71, 11.45) | <0.001 |

[1] Values are median (IQR) unless otherwise indicated.

[2] For continuous outcomes, this represents the Hodges-Lehman median of differences, with 95% confidence intervals calculated using normal approximation with continuity correction. Values < 0 indicate lower scores for CBT compared with control. For binary outcomes, this represents an odds ratio, estimated using logistic regression. Values > 1 indicate greater odds for CBT relative to control.

[3] For continuous outcomes, p values were estimated using the Wilcoxon rank-sum test. For binary outcomes, the Wald test was used to estimate p values.

Appendix 4. Worst-case sensitivity analysis for simulated ITT intervention effects on primary and secondary outcomes[1]

| | **CBT** | **Control** | **Comparison[2] (95% CI)** | ***P* value[3]** |
|---|---|---|---|---|
| Primary outcome | n = 79 | n = 74 | | |
| Endline aPHQ-9 | 3 (1, 4) | 8 (3, 10) | -4 (-5, -3) | <0.001 |
| Secondary outcomes | n = 79 | n = 74 | | |
| Change in aPHQ-9, endline - baseline | -9 (-10, -7) | -3 (-7, -1) | -4 (-6, -4) | <0.001 |
| Reduction in aPHQ-9 from baseline, n (%) | | | | |
| ≤ 3 | 7 (9) | 38 (51) | 0.09 (0.03, 0.22) | <0.001 |
| > 3 | 72 (91) | 36 (49) | 10.86 (4.65, 28.75) | <0.001 |
| > 5 | 66 (84) | 26 (35) | 9.37 (4.48, 20.75) | <0.001 |
| > 9 | 30 (38) | 4 (5) | 10.71 (3.92, 37.80) | <0.001 |
| > 50% | 68 (86) | 29 (39) | 9.59 (4.49, 21.98) | <0.001 |
| Endline aPHQ-9 < 5 | 59 (75) | 22 (30) | 6.97 (3.48, 14.50) | <0.001 |

[1] Values are median (IQR) unless otherwise indicated.

[2] For continuous outcomes, this represents the Hodges-Lehman median of differences, with 95% confidence intervals calculated using normal approximation with continuity correction. Values < 0 indicate lower scores for CBT compared with control. For binary outcomes, this represents an odds ratio, estimated using logistic regression. Values > 1 indicate greater odds for CBT relative to control.

[3] For continuous outcomes, p values were estimated using the Wilcoxon rank-sum test. For binary outcomes, the Wald test was used to estimate p values.

Appendix 5. Adjusted mITT intervention effects on primary and secondary outcomes[1]

| | **CBT** | **Control** | **Odds ratio[2] (95% CI)** | ***P* value[3]** |
|---|---|---|---|---|
| Primary outcome | n = 73 | n = 64 | | |
| Endline aPHQ-9 | 2 (1, 4) | 7 (3, 9) | 11.09 (5.43, 22.62) | <0.001 |
| Secondary outcomes | n = 73 | n = 64 | | |
| Change in aPHQ-9, endline - baseline | -9 (-11, -7) | -4 (-7, -2) | 11.36 (5.52, 23.37) | <0.001 |
| Reduction in aPHQ-9 from baseline, n (%) | | | | |
| ≤ 3 | 3 (4) | 29 (45) | 0.05 (0.01, 0.15) | <0.001 |
| > 3 | 72 (96) | 36 (55) | 20.80 (6.58, 93.08) | <0.001 |
| > 5 | 66 (88) | 26 (40) | 15.32 (6.17, 42.01) | <0.001 |
| > 9 | 30 (40) | 4 (6) | 19.28 (5.26, 102.69) | <0.001 |
| > 50% | 68 (91) | 29 (45) | 18.67 (7.10, 57.00) | <0.001 |
| Endline aPHQ-9 < 5 | 59 (79) | 22 (34) | 11.47 (4.89, 29.28) | <0.001 |
| Postnatal outcomes among antenatal enrollees by visit | | | | |
| 1.5 mo postnatal | n = 58 | n = 53 | | |
| aPHQ-9 score | 2 (0, 3) | 3 (0, 6) | 3.02 (1.50, 6.10) | 0.002 |
| aPHQ-9 < 5, n (%) | 49 (82) | 29 (55) | 4.36 (1.81, 11.19) | 0.001 |
| 3 mo postnatal | n = 57 | n = 49 | | |
| aPHQ-9 score | 2 (0, 4) | 4 (1, 7) | 2.95 (1.42, 6.14) | 0.004 |
| aPHQ-9 < 5, n (%) | 49 (83) | 26 (53) | 4.83 (1.94, 12.98) | 0.001 |
| 6 mo postnatal | n = 51 | n = 44 | | |
| aPHQ-9 score | 2 (0, 4) | 3 (0, 6) | 1.86 (0.89, 3.87) | 0.10 |
| aPHQ-9 < 5, n (%) | 40 (76) | 27 (61) | 2.09 (0.83, 5.46) | 0.12 |
| 9 mo postnatal | n = 46 | n = 36 | | |
| aPHQ-9 score | 0 (0, 3) | 2 (1, 5) | 3.86 (1.64, 9.07) | 0.002 |
| aPHQ-9 < 5, n (%) | 43 (90) | 26 (72) | 4.15 (1.17, 17.42) | 0.036 |

[1] Values are median (IQR) unless otherwise indicated. All analyses are adjusted for maternal age, education level, and baseline aPHQ-9 score. Two CBT participants and one control participant did not provide their age and were excluded from analyses. [2] For continuous outcomes, this represents an odds ratio for a lower aPHQ-9 score. Values > 1 indicate greater odds for lower scores for CBT compared with control. For binary outcomes, this represents an odds ratio, estimated using logistic regression. Values > 1 indicate greater odds for CBT relative to control.

[3] The Wald test was used to estimate p values for all outcomes.
